# Cost-effectiveness of mass drug administration with ivermectin against strongyloidiasis: a modelling study

**DOI:** 10.1101/2024.04.04.24305312

**Authors:** Luc E. Coffeng, Nathan C. Lo, Sake J. de Vlas

## Abstract

**Background:** Strongyloidiasis, caused by the parasitic intestinal worm *Strongyloides stercoralis*, infects hundreds of millions of people globally. Current school-based preventive chemotherapy (PC) programs that use benzimidazole derivatives (e.g., albendazole) against soil-transmitted helminths do not effectively treat strongyloidiasis, which requires treatment with ivermectin. We estimate the cost-effectiveness of mass drug administration with ivermectin for the control of strongyloidiasis.

**Methods:** We developed a mathematical model to simulate the population dynamics of *S. stercoralis* and the impact of school-based and community-wide PC across a range of epidemiological settings. We simulated 10-year PC programs with varying treatment coverages. We estimated a primary outcome of disability-adjusted life years (DALYs) averted by each PC strategy and calculate the programmatic cost (US$) of each strategy. We estimated cost-effectiveness by comparing strategies by their incremental cost-effectiveness ratios (US$/averted DALY) and expected loss curves.

**Findings:** The model found community-based PC was the most cost-effective strategy (≤600 US$ / DALY averted), despite costing approximately 5 times as much as school-based PC. Community-based PC targeted at ages 5 and above reduced infection levels close to 0% within 5 to 6 years. School-based PC was predicted to have very little impact. These results were robust across a range of epidemiologic settings above a measured prevalence of 2-5% in school age children.

**Interpretation:** Annual community-based PC is the most cost-effective public health strategy to control strongyloidiasis, being superior to school-based PC due to most of the infections and mortality occurring in adults. A baseline prevalence of 2% of infection in school age children, as measured by Baermann or stool culture, is a suitable minimum threshold for cost-effective implementation of community-based PC.

**Funding:** World Health Organization.

## Introduction

Strongyloidiasis is a neglected tropical disease caused by the soil-transmitted helminth (STH) *Strongyloides stercoralis* which is estimated to infect about 100–600 million people globally.^1,2^ After infection, newly established parasitic worms migrate via the lungs to the human gut where they cause local inflammation in the intestinal mucosa. Most infections are asymptomatic or mildly symptomatic, with chronic infection characterised by nonspecific symptoms such as abdominal pain, diarrhoea, hives, and anorexia.^3–5^ Acute strongyloidiasis may include symptoms of cough, dyspnoea, wheezing, haemoptysis, and vomiting.^3–5^ In endemic areas, the prevalence of strongyloidiasis can vary between only a few percent up to 40% in parts of South-East Asia.^5^ Typically, the prevalence of infection increases with age and then either stabilises at around 20–40 years^6,7^ or continues to rise.^8^ In immune-compromised individuals, loss of immune control can lead to hyperinfection with rapid multiplication of worm loads and disseminated infection with systemic sepsis and multi-organ failure, which is often fatal if untreated.^3^ The World Health Organization provides guidelines for the control of STH via regular deworming treatment and improved access to water, sanitation, and hygiene.^9^ However, these guidelines mostly focus on the control of other STH species (*Ascaris lumbricoides*, *Trichuris trichiura*, and hookworm species) and do not address control of strongyloidiasis and the challenges related to its complex life cycle, diagnosis, and treatment.

In contrast to other STH species, which are short-lived (up to three years) and can only be transmitted via the environment, strongyloidiasis is a life-long infection that self-renews through auto-infection.^3–5^ Transmission between humans is driven by fecal contamination of the environment and skin exposure to free-living infective filariform larvae (e.g., via walking barefoot), similar to hookworm transmission. Upon exposure, larvae penetrate the host skin and migrate via the lungs to the small intestine where they develop into patent adult female worms. After a pre-patent period of 3–4 weeks, the worms start producing eggs.^3^ Infections consist of solely female worms, which do not require presence of male worms to produce eggs (parthenogenesis).^3^ The eggs hatch when still in the human gut, where the emerged rhabditiform larvae can directly evolve into the filariform stage and re-infect the host (auto-infection).^3–5^ Egg production is strongly regulated by the host immune response, as evidenced by the phenomena of hyperinfection and disseminated infections in immunocompromised individuals.^3^ After hatching, rhabditiform larvae that are expelled into the environment can develop into one of two pathways.^3,5^ The first is the development into free-living infective filariform larvae that may survive in the environment for up to a few weeks. The second is the development into free-living rhabditiform adult male and female worms that live up to a few days and can reproduce sexually. Mated free-living female worms produce eggs, from which a new generation of rhabditiform larvae hatch which then develop into infective filariform larvae. The filariform larvae (from either developmental pathway) can infect new hosts via skin penetration. The offspring of free-living male and female worms cannot develop into free-living adult worms and must infect a human host for transmission to continue.^5^ As such, humans constitute the most important reservoir of infection and the environment is only a temporary reservoir.^5^

*S. stercoralis* infection cannot be reliably detected with diagnostic methods that are commonly used for other soil-transmitted helminths, such as microscopic examination of stool samples for detection of worm eggs (e.g., Kato-Katz).^10^ Even with diagnostic techniques that are optimised for strongyloidiasis, the sensitivity of a single stool-based diagnostic test (Baermann, Kogar Agar plate culture, or PCR) is in the order of 50% to 60%, while that of a single serological test is in the order of 65% (Bordier ELISA) to 80% (Strong Detect ELISA).^11^ As such, surveys to delineate STH-endemic areas typically underestimate the true prevalence of strongyloidiasis, especially when using stool microscopy. Combined with the lack of an established cut-off in terms of minimal prevalence of infection for initiating a control program, this poses a major challenge to decide where to implement strongylodiasis control measures.

In general, a main component of control strategies against STH is preventive chemotherapy (PC).^9^ The aim of PC is to bring down infection levels and transmission in endemic communities by regularly offering treatment to all eligible individuals in an endemic area, regardless of individuals’ infection status. However, current control programs against STH mainly treat school age children,^9^ using the benzimidazole derivatives albendazole or mebendazole which have limited activity against *S. stercoralis*.^12^ For treatment of strongyloidiasis, ivermectin is the drug of choice, which can be safely administered to individuals of age 5 (or 15 kg) and above.^12^ A single dose of ivermectin (200 μg/kg) cures 86% up to 95% of human infections.^13,14^ Repeated doses (up to four over two weeks) do not significantly improve this effect.^14^ *In vitro* studies suggest that ivermectin is active against both pre-patent and adult worms.^15^ Given the high efficacy and its successful and safe decades-long use in existing control programs against onchocerciasis and lymphatic filariasis, ivermectin is a prime candidate for PC against strongyloidiasis. In a recent meta-analysis of a highly heterogeneous set of population-based studies, single or repeated community-based PC with ivermectin was found to reduce prevalence of *S. stercoralis* infection by 82% on average.^16^ Also, infection levels in endemic communities do not seem to bounce back markedly between 3 and 9 months after a single round of PC.^17^ Still, the question remains in which epidemiological settings PC with ivermectin is a cost-efficient control strategy, and how this depends on which ages are targeted by PC. Targeting school age children (ages 5–15) via schools would be convenient as this strategy could be easily combined with existing STH control programs. On the other hand, community-wide PC (ages 5 and above) may be expected to have a larger health impact, but also to cost more.

Here, we estimate the public health impact and cost-effectiveness of PC programs with ivermectin for the control of strongyloidiasis. To this end, we developed a mathematical model to simulate the population dynamics of *S. stercoralis* and the impact of school-based and community-wide PC across a range of epidemiological settings. Cost-effectiveness and incremental cost-effectiveness ratios were estimated from a program perspective, using recent costing studies of school– and community-based PC programs against STH in Vietnam and the Philippines, as well as cost estimates for PC programs with ivermectin against onchocerciasis. Based on these findings, we formulate recommendations for where and how PC programs against strongyloidiasis should ideally be implemented.

## Methods

### Mathematical model structure and parameters

Given that patent *S. stercoralis* infection is self-replenishing and lifelong, transmission of *S. stercoralis* infection can be represented with a compartmental SEI model, where E (exposed) represents pre-patent infections and I (infectious) represent patent infections that may be transmitted to other hosts (Figure 1). Pre-patent infections were assumed to become patent in 3 to 4 weeks (Table 1).^3^ For patent infections, we assumed that without treatment, infections are indefinitely self-replenishing and therefore life-long. We further assumed that the effective rate at which individuals with patent infection excrete rhabditiform larvae is always the same, regardless of the female worm load, reflecting the strong regulation of worm fecundity by the host immune system. This approach was also convenient as very little is known about the distribution of *S. stercoralis* worm loads in human populations. However, a consequence of this assumption is that, in the model, the cure rate of treatment is the same for all treated individuals, while in other soil-transmitted helminthiases, this is known to decrease with higher worm loads.^18,19^

**Figure 1.**
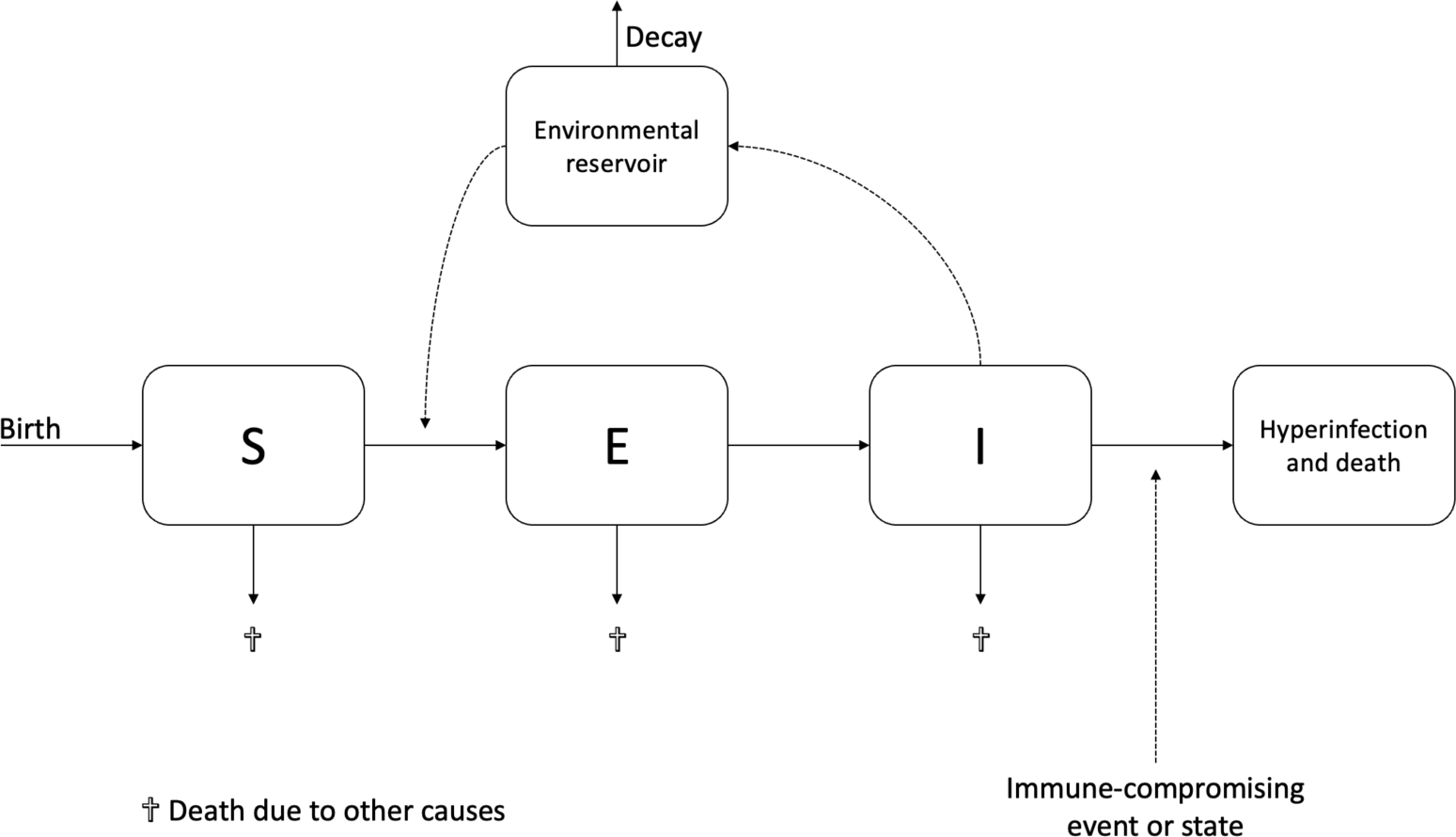
Schematic representation of an SEI model for *Strongyloides stercoralis* transmission. The model was implemented in a stochastic individual-based framework which explicitly includes age structure with a susceptible (S), exposed (E), and infectious (I) stage, along with an environmental reservoir. We account for risk of hyperinfection and death upon immunosuppression.

**Table 1.**
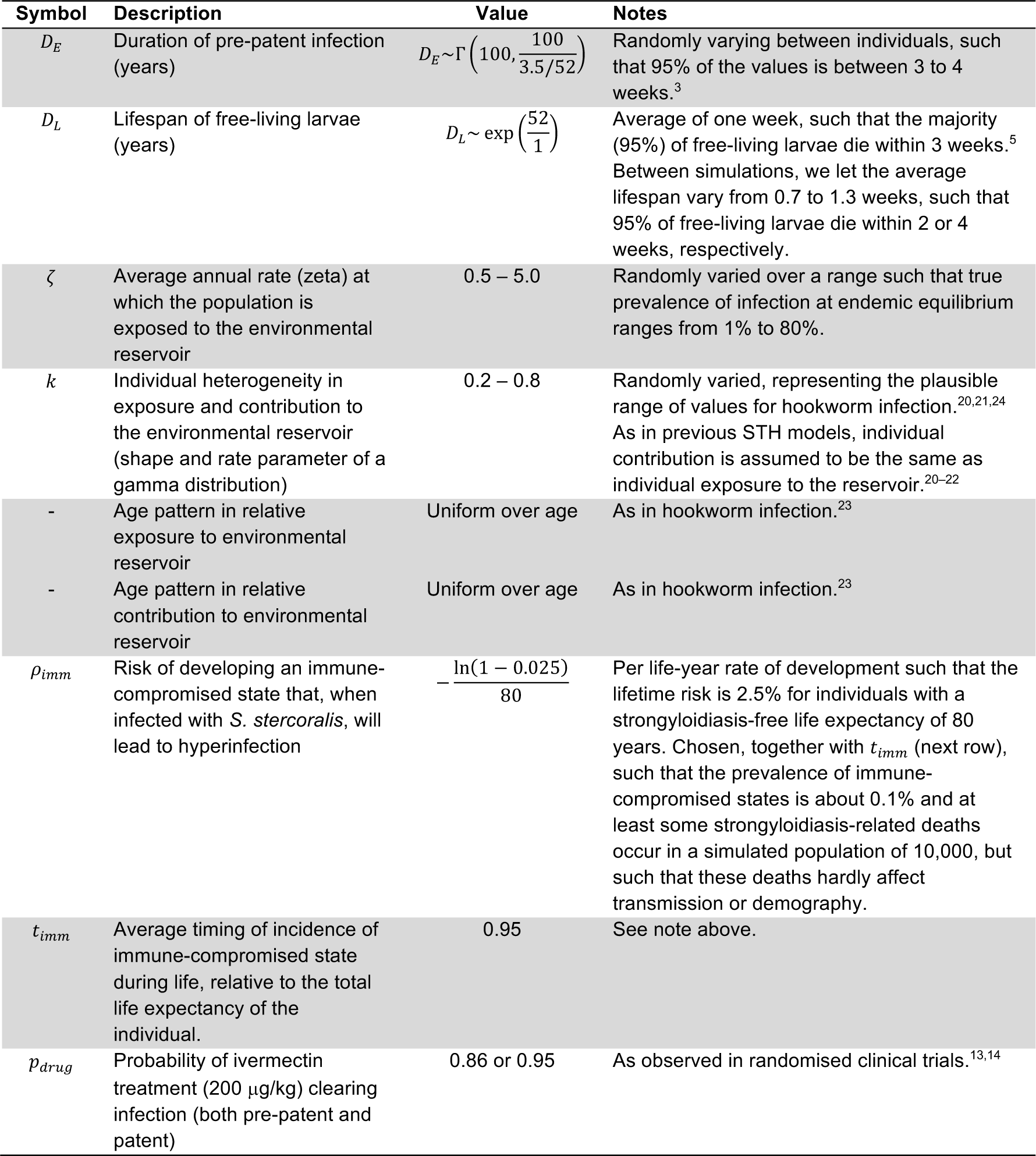
Parameter values for an SEI model of *Strongyloides stercoralis* transmission in humans.

As in previous transmission models for helminths,^20–22^ we included an environmental reservoir of infectious parasite life stages that decay over time (Figure 1). Given the relatively rapid dynamics of the environmental parasite life stages (days to weeks compared to years for the self-replenishing patent infections) (Table 1), we did not explicitly model free-living adult life stages or intermediate rhabditiform larval stages. Instead, as in other STH models,^20–22^ we assume that environmental contamination can directly lead to new transmission events. We adopted a uniform age pattern in exposure (i.e., all ages are exposed equally) from a transmission model for hookworm.^23^ In the past, patterns with higher exposure in adults have been proposed as well (which would favour community-based over school-based PC in terms of health impact),^20,21^ but these are not used here. We further assumed that individual exposure and contribution to the environmental reservoir of infection are highly heterogeneous due to variation in behaviour related to open defaecation and shoe-wearing.

As in previous modelling studies on other soil-transmitted helminthiases,^20,21^ we captured this heterogeneity with a gamma distribution for relative individual exposure and contribution to transmission, adopting parameter values for hookworm infection (Table 1).^20,21,24^ Although *S. stercoralis* can also cause patent infections in dogs,^5^ here we assume that no animal reservoir of infection is present.

With the model structure as described above, prevalence of infection in the model at endemic equilibrium is determined by three parameters: (1) the overall transmission rate, (2) the level of exposure heterogeneity, and (3) the survival of infective larvae in the environment. In addition, the level of exposure heterogeneity influences the curvature and saturation of the age pattern in infection levels. With higher heterogeneity (i.e., lower values of shape parameter *k*; Table 1), individuals on the higher end of the risk spectrum are infected earlier in life, creating a rise in prevalence at young ages which then tapers off at higher ages (when the lower-risk individuals are infected for the first time).

As *S. stercoralis* infections are lifelong and early-life infection may lead to hyperinfection and mortality much later in life, we explicitly captured the dynamics of human demography (birth, aging, and death) with a stochastic individual-based implementation of the SEI model, adopting mortality rates for sub-Saharan Africa for the year 2000.^25^ This life-course approach also means that we explicitly capture that individuals are at risk of being re-infected after no longer being targeted by PC (e.g., after leaving school in the context of a school-based PC program). Last, the simulated population was assumed to be closed, meaning that there was no migration in or out of the population. The simulated population size was kept stable by replacing every death by a birth. The resulting demographic structure of the population is presented in Supplemental Figure A, showing a high proportion of children in the population (favouring cost-effectiveness of school-based PC).

We modelled mortality due to *S. stercoralis* hyperinfection by letting a random fraction of simulated individuals develop an immune-compromised state. To determine whether an individual will develop an immune-compromised state during their lifetime, for each individual, we flip a coin weighted by the individual’s life expectancy (in absence of strongyloidiasis), such that people with longer life expectancies had a higher risk. The average incidence rate of immune-compromised states per year of life expectancy was set such that, for individuals with a life expectancy of 80 year, the cumulative incidence (i.e., lifetime risk) of this state was 2.5%. If the coin flip resulted in an immune-compromised state, this state was assumed to start when 95% of the individual’s natural lifespan had passed. We assumed that the individual would remain in that state until the end of their life; we did not explicitly model temporary immune-compromised states that may occur during a person’s life time. As a result of these assumptions, the overall prevalence of immune-compromised states in a simulated strongyloidiasis-free population was 0.1% and the age-specific prevalence only exceeded 0.2% for 60 and beyond (Supplemental Figure B). The average strongyloidiasis-free life expectancy of individuals that became immune-compromised was about 3.3 years (i.e., the years of life that could be potentially lost in case of strongyloidiasis infection). If at any point during the immune-compromised period, an individual had or contracted a patent infection, the individual was removed from the simulation and a “potential death” was recorded. Later on, when calculating the strongyloidiasis burden, we captured that some of these cases of hyperinfection are successfully diagnosed and treated before death (details below). We note that the data to inform the above assumptions are limited, although these assumptions were chosen such that potential deaths due to strongyloidiasis were so rare that they did not affect transmission or demography in any meaningful way, but were common enough for at least one strongyloidiasis-related death to occur in a simulated population of 100,000 over a 10-year period. This was important as we wanted to investigate to what extent uncertainty in the number of deaths and life years lost per death affected our cost-effectiveness estimates, which we did by scaling the number of model-predicted deaths with a constant (details below).

### Simulations and scenarios

First, we performed simulations for a closed population of 10,000 individuals across 1,500 random settings of transmission conditions such that the baseline true prevalence ranged between 0% and 80%. Transmission conditions were defined in terms of three parameters: the overall transmission rate, the level of variation in exposure between individuals (exposure heterogeneity), and the average lifespan of free-living infective larvae in the environment (for values, see Table 1). Parameter values for transmission conditions were sampled on a logarithmic scale, assuming a uniform distribution between the (logarithms of the) ranges listed in Table 1, using Latin hypercube sampling. Next, to assure that the simulated baseline (i.e., pre-treatment) infection levels reached equilibrium values, each simulation was run for 180 years with time steps of 1 day. The state of the population at the end of this simulation period and associated transmission parameters were saved into a databank to serve as a baseline setting from which to simulate forward. Simulations were categorised by baseline prevalence in school-aged children (age 5–15; prevalences 0-2, 2-5, 5-10, 10-15, 15-20, and ≥20), assuming the use of a coprological technique like Baermann or stool culture that detects infection with 50% sensitivity and 100% specificity.

For each of the 1,500 baseline settings, we predicted the impact of five scenarios: no PC, school-based PC at 80% or 95% coverage, and community-based PC at 65% or 80% coverage. These coverage values were informed by public health goals and common observed values. School-based PC was assumed to target children from age 5 to 15 and community-based PC was assumed to target all individuals of age 5 and above. Individual participation in repeated treatment rounds was assumed to be random. The treatment was assumed to cure pre-patent and patent infection with 86% probability.^14^ In sensitivity analyses, we investigated the impact of a cure rate of 95%^13^ and non-random participation (i.e., systematic non-participation) to PC.

Next, we combined sets of 10 simulations into populations of size 100,000, such that each population would contain at least on case of strongyloidiasis-related death. This was done by selecting 10 random simulations from the same endemicity category (prevalences in school age children of 0-2, 2-5, 5-10, 10-15, 15-20, and ≥20). This was repeated 1,000 times for each intervention strategy (N = 5) and endemicity category (N = 6), yielding 30,000 random sets (5 x 6 x 1,000) of 100,000 individuals.

### Health impact of PC strategies

The health impact of the different PC strategies was quantified in terms of disability-adjusted life years (DALYs) averted, where DALYs are the sum of Years of Life Lost (YLL) and Years Lived with Disability (YLD), weighted by the severity of disability. DALYs averted were calculated as the difference in DALYs lost between each intervention scenario and the reference scenario without PC. In sensitivity analyses, we also consider the health impact in terms of YLD averted only, which can be taken to represent settings with low risk of immune-compromising states or a degree of access to healthcare such that most cases of hyperinfection are diagnosed and treated before potential death.

### Years of life lost

We calculated the number of Years of Life Lost (YLL) assuming that upon hyperinfection, 20% (95%-CI: 10–30%) of cases survive by being diagnosed and treated via regular health care. For the untreated cases, we assume that they die and count the time they were planned to live (without strongyloidiasis) towards YLL. Our assumptions about the risk and timing of immune-compromising states (details above) implied that YLLs for an individual could at most be 5% of their strongyloidiasis-free life expectancy.

### Years lived with disability

The disease burden of prevalent cases of strongyloidiasis were calculated in terms of Years Lived with Disability (YLD), based on monthly model-predicted prevalences. We conservatively assumed that 25% (95%-CI: 20.9–29.4%) of all individuals with patent infections experience symptoms. This assumption was based on the observation that of people with detected infection based on Baermann or stool culture, 53% experience abdominal pain, 41% report diarrhoea, and 28% have urticaria or rash,^26^ combined with the fact that the sensitivity of Baermann or stool culture is in the order of 50%.^11,27,28^ With this approach, we ignore potential cases of undiagnosed infection who may yet have symptoms.

YLD were calculated as the total person-years of symptomatic infection times an average disability weight of 0.02 (95%-CI: 0.012–0.030), which is broadly consistent with previous calculations for other helminth infections.^26,29–31^ Some example disability weights of specific symptoms include mild diarrhoea (0.074), mild abdominopelvic problem (0.011), and symptomatic nematode infection (0.027).^30,31^ In addition, for each case of hyperinfection, we counted one person-month lived with a disability weight of 0.30 (95%-CI: 0.22–0.39), which is consistent with other severe infectious disease illnesses such as bloodstream infections, meningitis, and other disseminated infections ^30,31^.

### Cost and cost-effectiveness of PC strategies

For each deworming strategy, we calculated the cost from a control program perspective, accounting for cost of procurement and in-country distribution of ivermectin. We did not consider the cost of distribution of drugs to countries or productivity losses and (averted) costs related to hospital admissions. Also, we did not correct cost estimates for the year they were produced. All cost and cost-effectiveness analyses were performed assuming a time horizon of 10 years and a 3% discount rate for both costs and health effects. In sensitivity analyses, we investigated the impact of adopting a 5-year time horizon. In the next section, we detail the assumptions behind our cost and cost-effectiveness calculations. Wherever parameter uncertainty is indicated (95%-confidence intervals or 95%-CI), we propagated this in our calculations by using a randomly sampled value for each simulated population of 100,000.

### Cost of drug procurement

The cost per ivermectin treatment was assumed to be USD 0.10 for school age children and USD 0.30 for adults, with 95% probability that costs were in a range of ±10%. These values were based on the expected cost of generic ivermectin prequalified by WHO, as also recently used in a recent cost-effectiveness analysis by Buonfrate *et al*.^32^ In a sensitivity analysis, we also considered the option that drugs would be donated to control programs free of charge.

### Cost of delivery

For school-based PC implemented at 80% coverage and community-based PC at 65% coverage (which we consider acceptable coverage levels), we assumed that the cost of distribution of ivermectin was as estimated in a recent costing study of school-based PC and community-based PC against soil-transmitted helminths in Dak Lak province, Vietnam (Table 2).^33^ These estimates included the 10-year costs of training, community sensitisation, drug distribution (including capital costs, e.g., economic cost of using venues and class rooms), and monitoring and evaluation (deworming day inspections, reporting, parasitological surveys, and data management). We did not correct these estimates for the included cost of albendazole tablets (USD 0.03 per treatment), which contributed less than 5% to the total cost estimate.

**Table 2.**
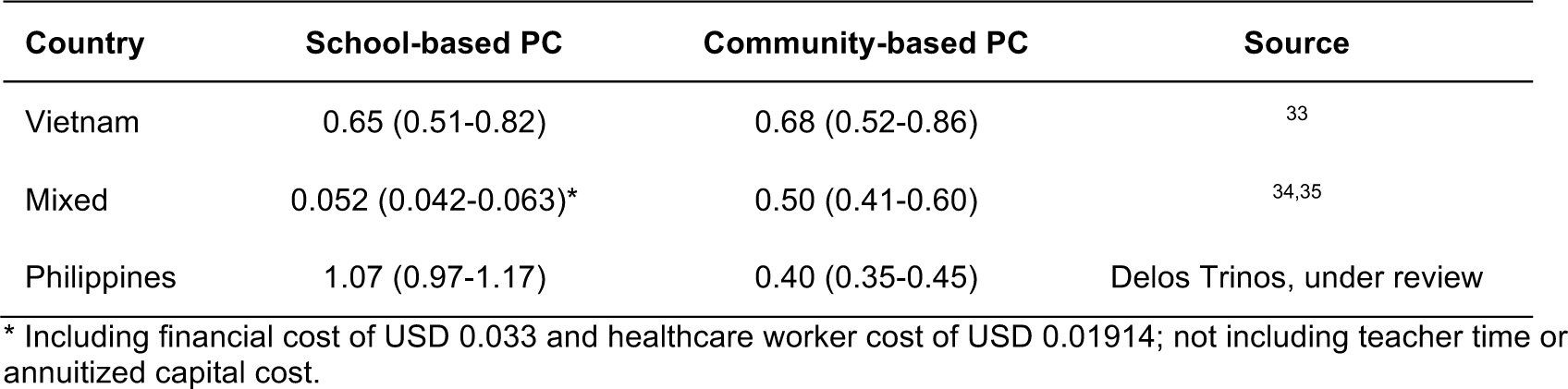
Assumed cost per treatment (USD) for delivery of school– and community-based PC, based on literature.

| Country | School-based PC | Community-based PC | Source |
| --- | --- | --- | --- |
| Vietnam | 0.65 (0.51-0.82) | 0.68 (0.52-0.86) | 33 |
| Mixed | 0.052 (0.042-0.063)* | 0.50 (0.41-0.60) | 34,35 |
| Philippines | 1.07 (0.97-1.17) | 0.40 (0.35-0.45) | Delos Trinos, under review |

\* Including financial cost of USD 0.033 and healthcare worker cost of USD 0.01914; not including teacher time or annuitized capital cost.

In a sensitivity analysis, we repeated all cost calculations adopting two alternative sets of assumptions (Table 2). The first was based on older estimates for PC distribution costs from literature from a mix of countries,^34,35^ for which the delivery cost per treatment of school-based PC was considerably lower than for community-based PC. As these estimates did not include uncertainty bounds, we assumed that the cost would be within a ±20% range around these values with 95% probability. The second set of alternative assumptions was based on a recent costing study for school– and community-based albendazole distribution in the Zamboanga Pensinsula of the Philippines (Delos Trinos *et al*, under review), where the delivery cost per treatment of school-based PC was higher than community-based PC (Table 2).

To capture that the distribution cost per treatment would probably be lower for control programs that achieved higher coverage (95% for school-based PC and 80% for community-based PC), we assumed that the increase in total delivery cost would on average be 50% (95%-CI: 31–69%) of the expected increase if the distribution cost per treatment were fixed.

For instance, scaling up from 80% to 95% coverage with a fixed distribution cost per treatment would imply an increase in total delivery cost of 18.8% (i.e., 95/80 − 1). Instead, we assumed that the total delivery cost would increase by 9.4% (95%-CI: 5.4–13.3%). Likewise, for school-based PC, we assumed that scaling up coverage from 65% to 80% would increase the total delivery cost by 11.5% (95%-CI: 6.7–16.4%) instead of the expected 23.1% (i.e., 80/65 − 1) in case of fixed delivery costs per treatment. To capture that for programs targeting larger populations, the delivery cost per treatment may drop considerably, we performed a sensitivity analysis assuming that the cost per treatment would be 75% lower for community-based PC^35^ and 50% lower for school-based PC,^36^ compared to the main analysis.

### Cost-effectiveness and incremental cost-effectiveness ratios (ICER)

For each interventions strategy and endemicity category, we calculated the cost-effectiveness in terms of cost (USD) per DALY averted. To demonstrate cost-effectiveness even in absence of (averted) strongyloidiasis-related mortality, the degree of which is a major uncertainty in our analysis, we also calculated the cost per YLD averted (i.e., assuming zero strongyloidiasis-related mortality). Based on the mean cost-effectiveness of each strategy, for each endemicity category we determined which strategies were on the cost-effectiveness frontier, using the non-iterative algorithm by Suen *et al*.^37^ Strategies that were not on the cost-effectiveness frontier were dominated, meaning that they either required higher costs and provided less benefit than other strategies (strictly dominated), or they did provide additional effects but at a higher cost per additional unit of effect than other strategies (extended or weakly dominated). For the strategies on the cost-effectiveness frontier, we calculated incremental cost-effectiveness ratios (ICERs), which expresses the cost per additional unit of effect gained when compared to the next-best strategy on the frontier. ICERs were calculated in two-fold, based either on DALYs or YLDs only. To investigate the sensitivity of cost-effectiveness on willingness to pay (WTP, in US dollar per DALY averted), we calculated and visualised expected loss curves,^38^ where the expected loss *L*_*s*_ of strategy *s*, given *N* repeated simulations, is defined as:

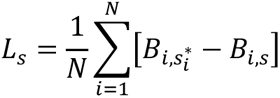

where 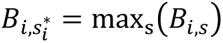, which is the net benefit of the optimal strategy (i.e., the highest net benefit) for the *i*-th simulations, denoted as 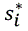. Here, the net benefit is simply product of the WTP and the number of DALYs averted by strategy *s*, minus the total cost of that strategy. As such, expected loss is a quantification of the expected foregone benefits of choosing a suboptimal strategy. For a given WTP, the most cost-effective strategy is the one that minimises the expected loss. Here, we considered a strategy cost-effective if it had the lowest expected loss for values of WTP ≤ US$1,085, which is the upper bound for low-income countries in terms of gross domestic product per capita.^39^

### Software

The stochastic individual-based transmission model was developed in R and is available as the open-source R package *strongysim* (v1.0.0) via https://gitlab.com/luccoffeng/strongysim. Simulations were run in R v4.2.1, using Rstudio v2023.03.0+386 as interface. Expected loss curves were produced with the decision analytic modeling package *dampack* v1.0.0 by Alarid-Escudero *et al*. (https://github.com/DARTH-git/dampack).^40^

### Adherence to reporting guidelines

In the supplemental information we describe how we adhered to the Guidelines for Accurate and Transparent Health Estimates Reporting (GATHER)^41^ and the principles of Policy-Relevant Items for reporting Models in Epidemiology of Neglected Tropical Diseases (PRIME-NTD).^42^

### Role of the funding source

The World Health Organization, which sponsored the study, provided the authors with access to the dossier and experts of the Guideline Development Group that was tasked with formulating a new guideline for control of strongyloidiasis. Other than that, the sponsor played no role in the interpretation or presentation of the data, the writing of the manuscript, or the decision to publish.

## Results

### Simulated baseline prevalences and age patterns

Based on the range of parameter values for transmission conditions in Table 1, we simulated stable baseline prevalences of *S. stercoralis* infection in the general population of between 0% and approximately 80% (as if measured by a perfect test). The baseline infection prevalence was determined mostly strongly determined by the overall transmission rate, less so by the exposure heterogeneity, and least by the average larval lifespan in the environment (Supplemental Figure C). The resulting distribution of prevalence of infection across the 1,500 baseline states is shown in Supplemental Figure D, where prevalence in school age children was lower than in the rest of the population. Model-predicted age patterns in infection prevalence rose up to approximately ages 10-15, after which they tapered off but continued to slowly rise with age (Figure 2). The steepness of the rise and tapering off depended on the overall endemicity level in the population. Given these age patterns in infection prevalence (Figure 2) and the age composition of the simulated population (Supplemental Figure A), at least 75% of infected cases were adults with higher percentages for settings with lower endemicity.

**Figure 2.**
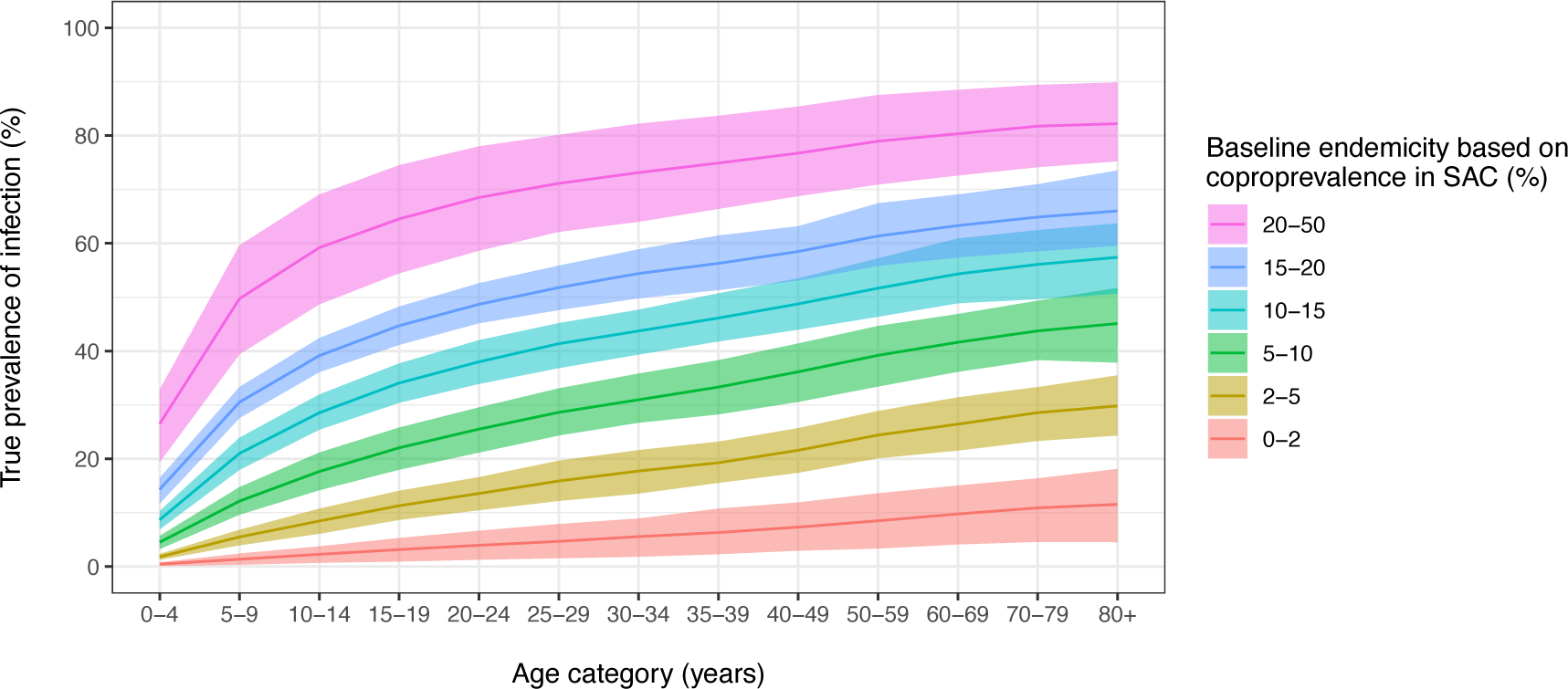
Model-predicted baseline age patterns in prevalence of infection, stratified by endemicity of *Strongyloides stercoralis*. Lines and ribbons represent the mean and 60%-confidence interval of simulated populations of 10,000 people. Age patterns are stratified by baseline endemicity (colours) as measured in school age children (SAC) with a coprological test (50% sensitivity).

### Predicted trends in infection prevalence

The model-predicted impact of PC strongly depended on the type of implementation (Figure 3). School-based PC reduced infection levels in the population to some extent over 10 years, but not by more than 12 percentage points for settings with the highest infection levels (pink). Even for the lowest endemic settings (red), the model predicted that school-based PC would only marginally impact infection levels, which is directly related to most infections occurring in adults. Increasing the coverage of school-based PC from 80% to 95% only marginally improved the impact of PC on infection levels. In contrast, for all but the highest endemic settings, community-based PC led to a reduction of infection levels close to zero within 5 years, even when coverage of the target population (age 5 and above) was 65%. For the highest endemic settings, increasing the coverage to 80% reduced infection levels close to zero within 5 years as well.

**Figure 3.**
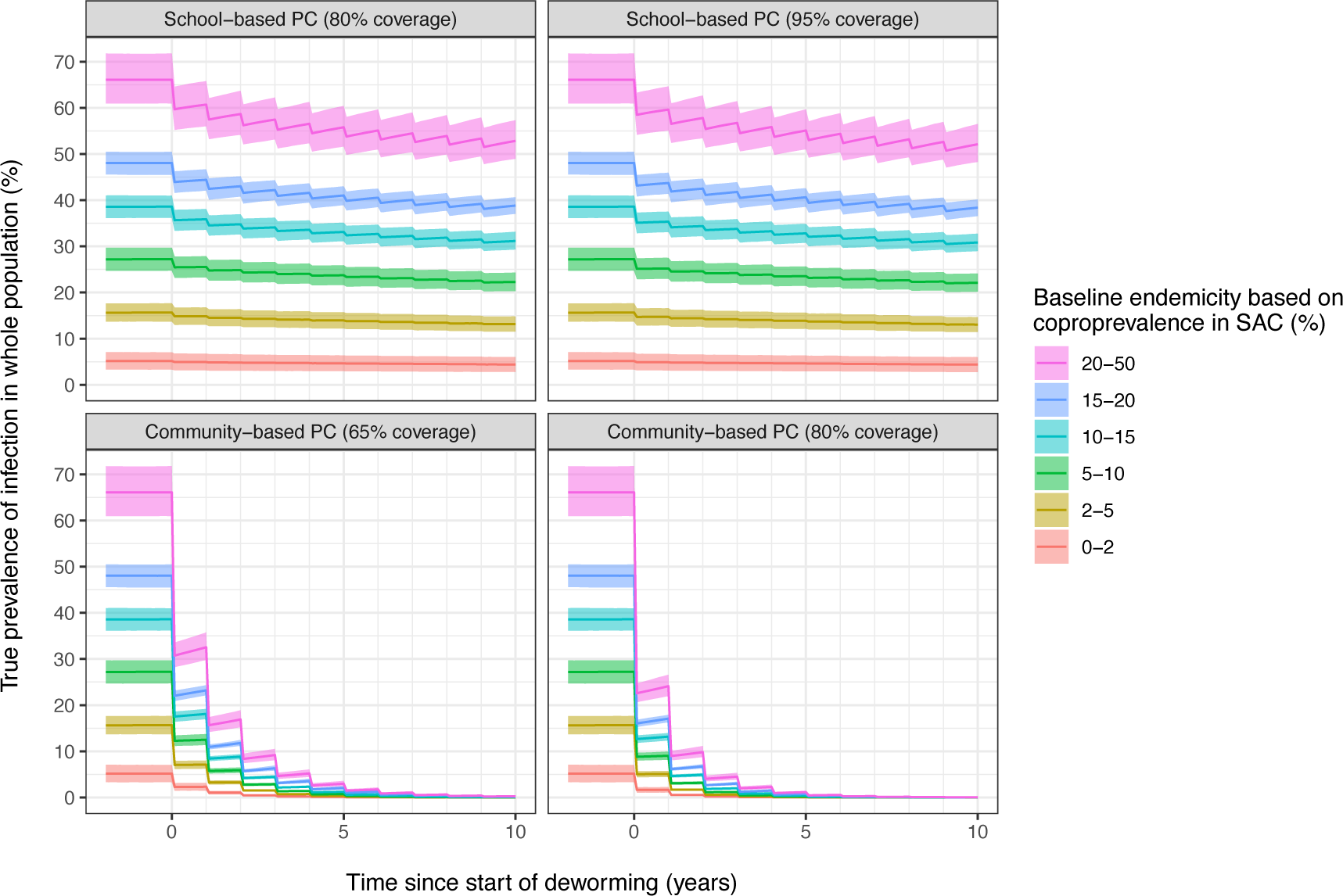
Model-predicted impact of different preventive chemotherapy (PC) strategies on true prevalence of *Strongyloides stercoralis* in a population of 100,000 people. Trends are stratified by baseline endemicity (colours) as measured in school age children (SAC) with a coprological test (50% sensitivity, 100% specificity). Lines and ribbons represent the mean and 95%-confidence interval of predicted values per endemicity category.

### Impact of preventive chemotherapy on disease burden

Based on the predicted trends in infection and hyperinfections, we calculated DALYs as the sum of YLDs (based on infection prevalence) and YLLs (based on hyperinfections) from the start of PC until 10 years after. We note that the predicted number of hyperinfections is inherently subject to a high degree of uncertainty due to limited data regarding the prevalence and age pattern of immune-compromising conditions and context-dependent access to healthcare. For a population of 100,000 without PC, the 10-year burden of strongyloidiasis varied from 304 DALYs lost for the lowest endemic setting, up to 3,553 DALYs lost for the highest endemic settings (Supplemental Table A). For these settings, YLDs contributed about 2.5–4 times as much to DALYs as YLL, with YLD contributing relatively more in highly endemic settings due to higher infection prevalence in younger people who are at low risk of hyperinfection. Compared to the scenario without PC, school-based PC reduced the number of DALYs lost by 6% (lowest endemic settings) to 14% (highest endemic settings), and mostly via a reduction in YLDs lost (Table 3). In contrast, community-based PC resulting in a marked decrease of both YLLs and YLDs, reducing DALYs by 85% (PC at 65% coverage) up to 89% (PC at 80% coverage) across all endemicity levels.

**Table 3.** Model-predicted averted burden during 10 years of preventive chemotherapy against strongyloidiasis per 100,000 population. Point estimates represent medians over repeated simulated populations. Numbers in brackets represent the central 95%-confidence intervals that capture uncertainty about disability weights, the fraction of the hyperinfections that are prevented from dying via regular routine healthcare, and stochastic variation in a population of 100,000 people. All burden estimates include an annual discounting rate of 3%.

| Baseline prevalence in SAC (%)* | PC Strategy | PC Coverage | YLDs averted |  | YLLs averted ** |  | DALYs averted |  |
| --- | --- | --- | --- | --- | --- | --- | --- | --- |
| 0-2 | School-based PC | 80% | 20 | (10-37) | -1 | (-10-7) | 19 | (6-39) |
|  |  | 95% | 21 | (10-40) | 0 | (-8-9) | 21 | (7-42) |
|  | Community-based PC | 65% | 197 | (98-352) | 60 | (29-100) | 258 | (142-428) |
|  |  | 80% | 205 | (102-366) | 62 | (30-105) | 267 | (146-445) |
| 2-5 | School-based PC | 80% | 75 | (44-122) | 0 | (-12-13) | 76 | (42-125) |
|  |  | 95% | 79 | (47-128) | 0 | (-13-12) | 80 | (45-132) |
|  | Community-based PC | 65% | 602 | (360-972) | 177 | (130-236) | 775 | (538-1,164) |
|  |  | 80% | 628 | (375-1,010) | 185 | (135-246) | 809 | (562-1,209) |
| 5-10 | School-based PC | 80% | 151 | (87-233) | 2 | (-10-15) | 155 | (89-239) |
|  |  | 95% | 160 | (92-247) | 3 | (-10-16) | 163 | (94-255) |
|  | Community-based PC | 65% | 1,050 | (610-1,625) | 267 | (207-342) | 1,316 | (879-1,911) |
|  |  | 80% | 1,097 | (637-1,694) | 278 | (215-354) | 1,371 | (912-1,992) |
| 10-15 | School-based PC | 80% | 235 | (137-368) | 3 | (-8-14) | 238 | (140-368) |
|  |  | 95% | 250 | (146-393) | 3 | (-8-15) | 252 | (149-393) |
|  | Community-based PC | 65% | 1,478 | (861-2,314) | 337 | (270-419) | 1,813 | (1,206-2,642) |
|  |  | 80% | 1,545 | (900-2,425) | 352 | (284-434) | 1,893 | (1,254-2,769) |
| 15-20 | School-based PC | 80% | 305 | (177-470) | 2 | (-8-12) | 306 | (178-476) |
|  |  | 95% | 325 | (191-506) | 2 | (-9-11) | 326 | (193-506) |
|  | Community-based PC | 65% | 1,822 | (1,061-2,822) | 388 | (312-481) | 2,210 | (1,462-3,213) |
|  |  | 80% | 1,910 | (1,111-2,962) | 409 | (331-504) | 2,318 | (1,535-3,377) |
| 20-50 | School-based PC | 80% | 454 | (261-693) | 3 | (-5-12) | 457 | (264-692) |
|  |  | 95% | 489 | (281-749) | 3 | (-6-11) | 490 | (284-746) |
|  | Community-based PC | 65% | 2,511 | (1,449-3,800) | 496 | (408-608) | 3,002 | (1,926-4,307) |
|  |  | 80% | 2,637 | (1,522-3,984) | 523 | (428-637) | 3,156 | (2,027-4,535) |

\* Baseline prevalence in school age children (SAC), assuming 50% sensitivity and 100% specificity of the diagnostic test used.
\*\* Negative values are due to stochastic variation in simulations.
PC = preventive chemotherapy; YLL = Years of Life Lost; YLD = Years Lived with Disability; DALY = Disability-Adjusted Life-Years.

### Cost and cost-effectiveness

The estimated total cost of community-based PC was roughly five times that of school-based PC (Table 4). The cost-effectiveness of school-based PC ranged from 4,716 USD per DALY averted for lowest endemic settings to 198 USD per DALY averted for the highest endemic settings (Table 5). For community-based PC, the cost-effectiveness was always more favourable than for school-based PC and ranged from 2,063 USD per DALY averted for the lowest endemic areas to 161 USD per DALY averted for the highest endemic areas. For both school– and community-based PC, improved coverage levels (95% and 80%, respectively) slightly increased the cost per DALY averted. In terms of the incremental cost-effectiveness ratio (ICER), school-based PC was always dominated by community-based PC (Table 5). This means that although school-based PC averted DALYs, it did so at a higher cost per DALY averted than community-based PC. Therefore, community-based PC at 65% of the target population was always the first strategy on the cost-effectiveness frontier (after “No PC”), and served as the reference for calculation of the ICER of community-based PC at 80% coverage. The ICER of community-based PC at 80% coverage was always considerably higher (roughly 3 to 4-fold) than that for 65% coverage, meaning that the additional DALYs that 80% coverage averted in surplus of those that would be averted by 65% coverage were relatively more expensive. Further, ICERs for community-based PC strongly depended on the baseline endemicity. For example, for 80% coverage, ICERs ranged from 7,034 USD per additional DALY averted in low endemic settings to 473 USD per additional DALY averted. ICERs based on YLDs averted only (i.e., assuming absence of strongyloidiasis-related mortality) were higher than those based on DALYs, although they were of the same order of magnitude and showed the same patterns in terms of school-based PC being dominated (Supplemental Table B).

**Table 4.** Estimated cost of 10 years of school-based and community-based preventive chemotherapy (PC) in a population of 100,000. Costs are expressed in thousands of US dollars. Cost of distribution of ivermectin was assumed to be as estimated for preventive chemotherapy against soil-transmitted helminths in Dak Lak province, Vietnam.^33^

| PC Strategy | PC Coverage | Total cost (USD x 1,000) |  |
| --- | --- | --- | --- |
| School-based PC | 80% | 90 | (72-110) |
|  | 95% | 99 | (80-123) |
| Community-based PC | 65% | 481 | (397-577) |
|  | 80% | 552 | (460-662) |

**Table 5.**
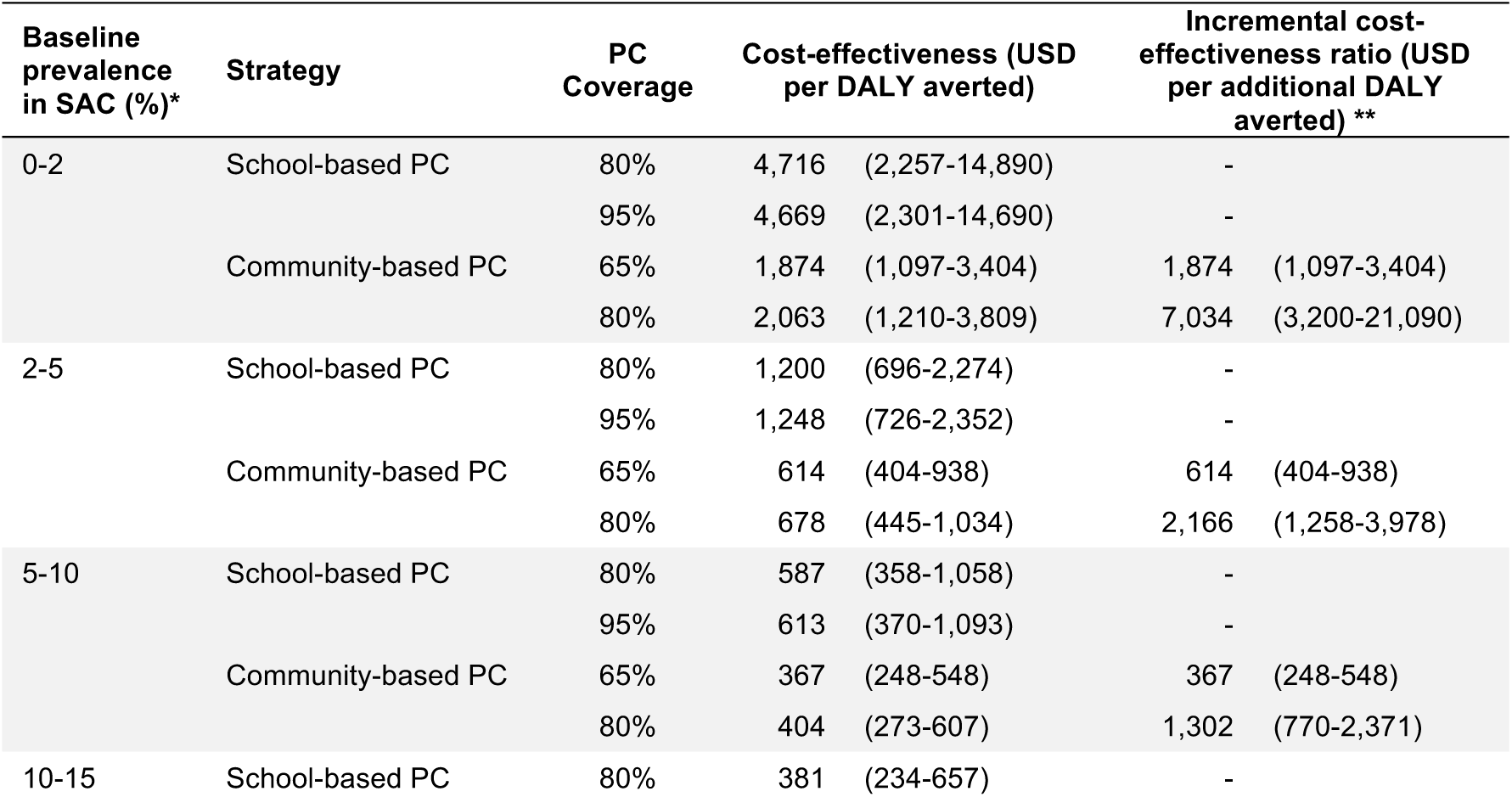
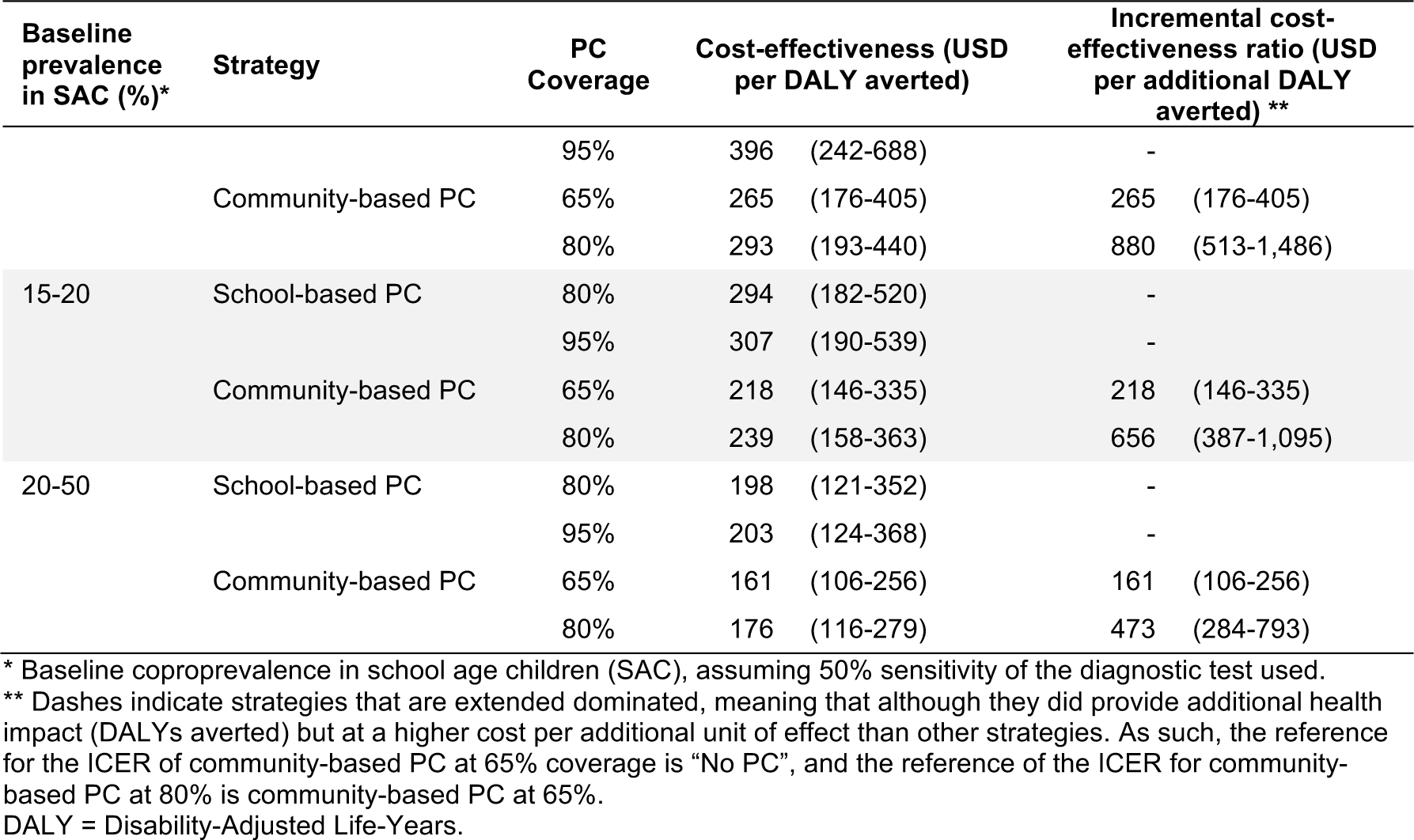
Cost-effectiveness and incremental cost-effectiveness ratios of different 10-year preventive chemotherapy (PC) strategies, stratified by endemicity. Note that costs-effectiveness and ICER estimates are expressed in US dollars (not thousands). Cost of distribution of ivermectin was assumed to be as estimated for preventive chemotherapy against soil-transmitted helminths in Dak Lak province, Vietnam.^33^ Point estimates represent medians over repeated simulated populations of 100,000 people. Numbers in brackets represent the central 95%-confidence intervals that capture uncertainty about costs, disability weights, the fraction of the hyperinfections that are prevented from dying via regular routine healthcare, and stochastic variation in a population of 100,000 people.

Because the choice of most cost-effective strategy depends on the WTP per DALY averted, we calculated the expected loss of each PC strategy as a function of the WTP. Here, the expected loss is a quantification of the expected foregone benefits of choosing a suboptimal strategy. For a given WTP, the strategy with the lowest expected loss (i.e., the lowest foregone benefits) is the most cost-effective strategy. For settings with prevalence of infection <2% (in school age children, as measured by a 50% sensitive coprological test), no PC was the most cost-effective strategy up to a WTP of 1,850 USD per DALY averted; for higher WTP, community-based PC at 65% coverage was more cost-effective (Figure 4). For settings with a baseline prevalence of 2-5%, community-based PC at 65% coverage was the most cost-effective for a WTP of at least 600 USD per DALY averted, and increasing coverage to 80% would be the most cost-effective for WTP slightly over 2,000 USD per DALY averted. For higher baseline endemicity levels, community-based PC was the most cost-effective for even WTP under USD 600, and higher PC coverage (80% instead of 65%) became increasingly cost-effective.

**Figure 4.**
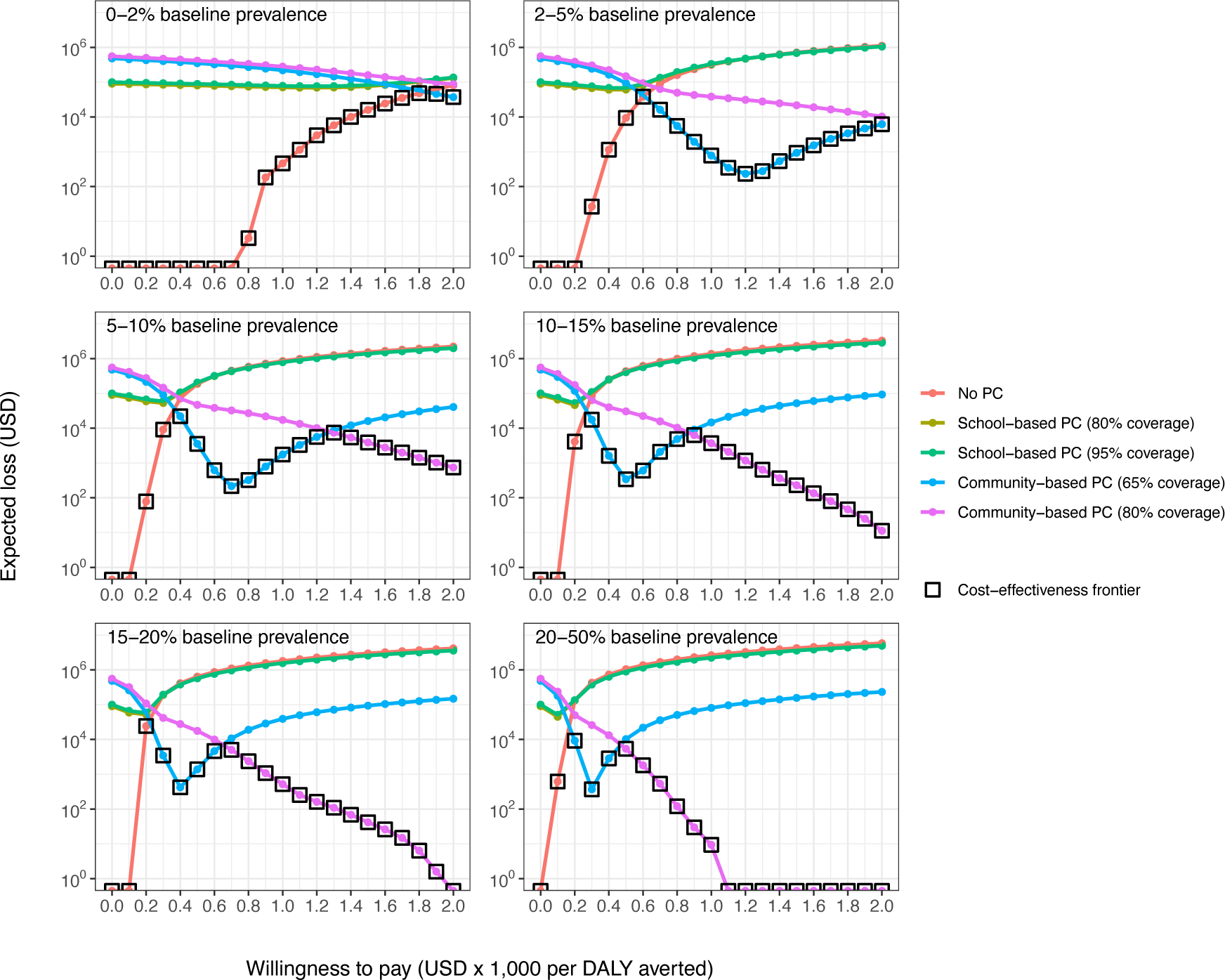
Expected loss in US dollars as a function of willingness to pay (WTP) per DALY averted in a population of 100,000 individuals. For a given WTP, the most cost-effective strategy is the one that minimises the expected loss (indicated with black open squares). Cost of distribution of ivermectin was assumed to be as estimated for preventive chemotherapy against soil-transmitted helminths in Dak Lak province, Vietnam.^33^ The line for school-based PC at 80% coverage is largely hidden behind the line for school-based PC at 95% coverage. Panels represent settings with different baseline infection prevalence in school age children, as measured with a coprological test (assuming 50% sensitivity and 100% specificity). Bullets on the horizontal axis indicate situations where the expected loss was zero, meaning that a strategy always produced the highest net benefits and was the most cost-effective across all simulated populations and across the range of uncertainty about cost and health impact.

### Sensitivity analyses

Our first sensitivity analysis addresses uncertainty in YLL, which is important given the major uncertainty about (1) age patterns in the incidence and prevalence of immune-compromised state, which likely vary between contexts; (2) the remaining life expectancy and quality of life after incidence of immune-compromised states (in absence of strongyloidiasis), (3) the fatality rate among cases of hyperinfection, which may again vary between contexts, depending on access to diagnosis and treatment. For these reasons, we also calculated the expected loss of PC strategies based on averted YLD only. Based on this, community-based PC at 65% coverage still was cost-effective for settings with at least 2% baseline prevalence of infection in school age children and a WTP of 800 USD per YLD averted (in Table 6).

**Table 6.** Sensitivity analysis for the minimal willingness to pay (in USD, rounded to nearest multiple of 50) per DALY averted for annual community-based PC (implemented at 65% coverage) to be cost-effective. Baseline infection prevalence is assumed to be measured with a 50% sensitive diagnostic test.

| Assumption | Baseline prevalence $\geq 2\%$ | Baseline prevalence $< 2\%$ |
| --- | --- | --- |
| Main analysis | 600 | 1,850 |
| Cost-effectiveness based on YLDs only | 800 | >2,000 |
| Higher impact of ivermectin (cure rate of 95% instead of 86%) | 600 | 1,800 |
| Time horizon of 5 years instead of 10 years | 800 | >2,000 |
| PC implemented for 5 years (i.e., 6 rounds) instead of 10 years | 350 | 1,000 |
| Systematic non-participation to community-based PC (7.5% never treated after 5 rounds) | 650 | 1,950 |
| Delivery cost of USD 0.50 per treatment for community-based PC (instead of USD 0.68) and USD 0.05 per treatment for school-based PC (instead of USD 0.65) | 550 | 1,550 |
| Delivery cost of USD 0.40 per treatment for community-based PC (instead of USD 0.68) and USD 1.07 per treatment for school-based PC (instead of USD 0.65) | 450 | 1,300 |
| National program perspective, ignoring the economic cost of donated drugs | 450 | 1,300 |
| Economies of scale for large programs ($>100,000$ population), assuming 75% lower delivery cost for community-based PC and 50% lower for school-based PC | 500 | 1,500 |
| No (additional) delivery cost for school-based PC due to existing program | 600 | 1,850 |

If ivermectin was assumed to be more effective than in our main analysis (cure rate of 95% instead of 86%), predicted trends in infection would on change marginally. School-based PC would still have relatively little impact, and the already large impact of community-based PC would increase only marginally. Likewise, the expected loss of each strategy remained very similar to the main analysis, with community-based PC being cost-effective from a WTP of USD 600 per DALY averted and baseline prevalence of 2%.

As most of the impact of community-based PC on infection levels was predicted to occur in the first 5 years of a PC program, we also considered a time horizon of 5 years instead of 10. Here, the minimal WTP for community-based PC to be cost-effective increased from USD 600 to USD 800 per DALY averted, compared to the main analysis. In addition, increasing coverage of community-based PC from 65% to 80% was cost-effective more often than in the main analysis (for lower WTP and lower endemicity levels). We further considered the implementation of only 5 years of PC (i.e., 6 treatment rounds), while keeping the time horizon at 10 years. This allowed us to account for potential elimination or bounce-back of infection levels after stopping PC. As bounce-back of infection levels after the last PC round was relatively slow, community-based PC was still the most cost-effective strategy for settings with a WTP of at least 350 USD per DALY averted and a baseline prevalence of 2%. Even for settings with a baseline prevalence <2%, community-based PC at 65% coverage was cost-effectiveness for a WTP of at least 1,000 USD per DALY averted.

As achieving high coverage is more challenging for community-based than school-based PC, we also considered the possibility of systematic non-participation in PC being considerable in community-based PC. We therefore assumed that after 5 rounds of community-based PC, 7.5% of the population (that was eligible for treatment all those 5 rounds) never took treatment (instead of <0.5% when assuming random participation). Under these assumptions, community-based PC still reduced infection prevalence to 0-5% within 5 years, which was not as close to zero percent as in the main analysis. Nevertheless, community-based PC was still the most cost-effective strategy for a WTP of at least USD 650 per DALY averted in settings with baseline prevalence of at least 2%.

Last, and importantly, we consider several alternative assumptions about the cost of PC. First, we adopted estimates of the delivery cost from literature that are similar for community-based PC (USD 0.50 per treatment instead of USD 0.68) but substantially lower for school-based PC than in our main analysis (USD 0.0514 per treatment instead of USD 0.65). Based on these cost estimates, school-based PC was no longer dominated and became a viable cost-effective strategy, but only for a narrow range of WTP in the order of USD 250–500 per DALY averted (baseline prevalence of 2%) to USD 100–150 (highest endemic areas). For higher WTP, community-based PC was still the most cost-effective strategy. Second, we adopted cost estimates from the Zamboanga Peninsula (the Philippines), for which the delivery cost per treatment was lower for community-based PC (USD 0.40 instead of USD 0.68) but higher for school-based PC (USD 1.07 instead of USD 0.65). With these cost estimates, community-based PC was the most cost-effective strategy for WTP of USD 450 per DALY averted in baseline prevalence of at least 2%. And for settings with a baseline prevalence <2%, community-based PC was cost-effective if the WTP was at least USD 1,300 per DALY averted. When considering the cost from a national program perspective, ignoring the economic cost of potentially donated drugs, community-based PC was also the most cost-effective strategy for WTP of USD 450 per DALY. Further, we considered that the targeted population was larger than 100,000 population, assuming that this would reduce the delivery cost per treatment by 75% for community-based PC and by 50% for school-based PC due to economies of scale. Here, community-based PC was the cost-effective strategy for WTP of at least USD 500 per DALY averted for baseline prevalences of at least 2%, and WTP of at least USD 1,500 per DALY averted for baseline prevalences <2%. Last, we considered the possibility that school-based PC could be delivered via an existing program, considering only the cost of the drug and assuming zero additional delivery costs. Given the small impact of school-based PC on overall infection levels, this assumption did not change conclusions about cost-effectiveness or the minimum WTP for community-based PC to be the most cost-effective option (Table 6).

### Optimal number of community-based PC rounds

Given that sensitivity analyses confirmed that community-based PC always dominates school-based PC in terms of cost-effectiveness, we briefly considered the optimal number of rounds of annual community-based PC. To this end, we simulated 1 up to 10 PC rounds at 65% or 80% coverage, while maintaining the rest of the assumptions as in the main analysis, including the 10-year time horizon. Given that annual community-based PC quickly reduces the prevalence of strongyloidiasis infection and given that prevalence bounces back only slowly after PC (Figure 3), implementing more than 5 PC rounds only averted a marginal number of additional DALYs (Supplemental Figure E) while adding cost for implementation for PC.

We determined the optimal number of rounds of community-based PC based on ICERs of increasing number of PC rounds, assuming that the cost per treatment does not change with the duration of the program. In Figure 5, we show that 3 to 6 rounds of community-based PC are cost-effective for settings in low-income countries (bottom dashed line) with a baseline prevalence of infection in SAC of ≥2%. For lower middle-income countries (top dashed line), a strategy of up to 7 annual PC rounds was predicted to be cost-effective, but only in very highly endemic settings (baseline prevalence of ≥15% combined with PC coverage of 65%). We further note that with higher coverage (80% vs. 65%), the optimal number of rounds of community-based PC was decreased by at most 1. When basing ICERs only on disability and not mortality (i.e., YLD averted), the optimal number of rounds was either the same or one round less (Supplemental Figure F).

**Figure 5.**
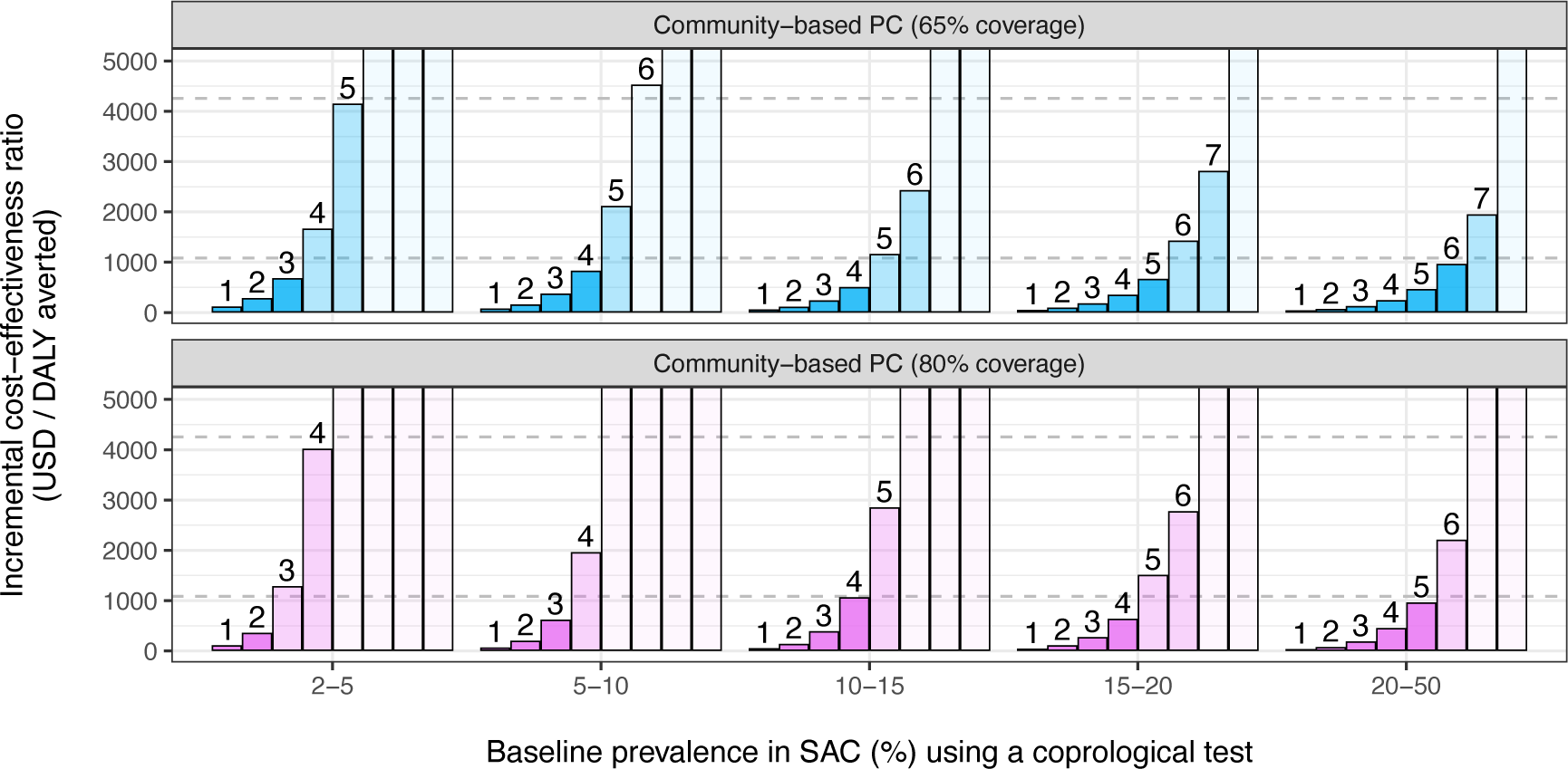
Incremental cost-effectiveness ratios (ICERs) for increasing number of annual rounds of community-based preventive chemotherapy (PC). The number over each bar indicates the number of PC rounds, where the colour of bars indicates whether the ICER for that number of PC rounds was under the average gross domestic product (GDP) for low-income countries (US$1,085, lower dashed line) or under the average GDP of lower middle-income countries (US$4,255, upper dashed line), or over.^39^ ICERs greater than US$5,000 are capped at the top of each panel. Note that ICERs may be underestimated for the shortest PC program durations. This is because an average cost per treatment (calculated for a 10-year period) was applied to all PC program durations, not considering a potential concentration of costs at the start of PC programs due to initial start-up costs. This also means that the ICERs for longer program durations over-estimated somewhat.

## Discussion

In this modelling study, we estimated the public health impact and cost-effectiveness of PC with ivermectin for control of strongyloidiasis, concluding that community-wide PC is likely to have a large impact and be cost-effective. Assuming that untreated *S. stercoralis* infection is life-long, we predict that its transmission dynamics and the bounce-back in population infection levels after PC are slow. As the majority of infections occur in adults, school-based PC was predicted to have very little impact on the burden of strongyloidiasis, in contrast to community-based PC targeted at ages 5 and above. For a 10-year PC program, the most cost-effective strategy was annual community-based PC in settings with at least 2% baseline prevalence in school age children (measured with a 50% sensitive diagnostic test). As most of the health impact was predicted to be achieved in the first five years of PC, 3 to 6 rounds of annual PC implement at ≥65% coverage can be safely recommended, with the option of up to 7 for highest endemic areas (baseline prevalence ≥15%). Implementing community-based PC at 80% instead of 65% coverage may reduce the optimal number of PC rounds by one, potentially saving some costs. In a wide range of sensitivity analyses for assumptions about strongyloidiasis-related mortality, the cost of PC, drug efficacy, systematic non-participation to PC, duration of PC, and the time horizon considered, community-based PC remained the most cost-effective strategy. We further found that the cost-effectiveness of community-based PC increases considerably with baseline endemicity, meaning that more highly endemic areas should be prioritised for PC.

Our estimates suggest that community-based PC against strongyloidiasis is within the same order of cost-effectiveness (USD 100–1000 per DALY averted) as, for instance, PC against other soil-transmitted helminths and schistosomiasis, treatment of colorectal cancer (in low-income countries), non-price interventions for tobacco, treatment of tuberculosis with second-line drugs (in middle-income countries), and scale-up of anti-retroviral therapy for HIV infection to all infected persons (in South Africa).^43^ In addition, it may even be possible to stop community-based PC within as little as 5 or 6 years if and when it leads to interruption of transmission, especially when combined with improved access to and uptake of water, hygiene and sanitation (WASH).^44^ In contrast, to maintain the health impact of school-based PC, it would have to be continued indefinitely or transmission would have to be reduced via WASH.

Our findings contrast those found in a recent cost-effectiveness study which suggested that school-based PC was more cost-effective against strongyloidiasis than community-based PC.^32^ The authors performed a Markov-model-like analysis and estimated that the cost per recovered (i.e., infection-free) person was US$1.97 for school-based PC and US$3.43 for community-based PC, and that the cost per averted death for a 10-year time horizon was US$288 (school-based PC) or US$969 for (community-based PC). The key differences in this study that explain the difference in findings include their assumptions that the risk of hyperinfection and death was the same for all ages, whereas this is probably low in children and higher in the elderly. As such, their estimates of cost-effectiveness are likely to be overly optimistic for school-based PC and pessimistic for community-based PC. In addition, in their Markov-model-like analysis, the authors assumed a static infection model; after each round, the annual incidence of new infections was assumed to be 50% of the baseline prevalence, which implies a degree of bounce-back that is much higher than suggested by our dynamic transmission model. As a result, the authors’ model-predicted impact of PC on infection levels was lower than predicted by our transmission model, which probably led to an overestimation of the cost per recovered person in their analysis. The authors further assumed that each year, 0.4% of all infected cases would develop severe strongyloidiasis of which 64% would die (i.e., 0.26% of all infections each year). This number was in the same order of magnitude as our predictions for the number of strongyloidiasis-related deaths per person-year with infection (across the whole population and 10-year time period), which, when translated to percent per infected case per year, ranged from 0.29% for the lowest endemic settings (where infection occurs more so in adults) to 0.19% for the highest endemic settings (in which individual are infected earlier in life on average).

In another recent mathematical modelling study of strongyloidiasis,^45^ others explicitly modelled the complexities of the parasite life cycle in the environment (mating of free-living female and male worms) as well as the auto-infection process and regulation of female worm egg production by the host immune response. In comparison, our model here is simpler and has fewer parameters and effectively assumes that (1) the mating process of free-living adult worms amplifies the size and longevity (somewhat) of the environmental reservoir of infection by a constant and can therefore be left out and absorbed into the central parameter for overall intensity of transmission (ζ); (2) the host immune response and the resulting regulation of female worm fecundity reaches it maximum in negligible little time (weeks to months) compared to the duration of untreated infection (years to lifetime) and can therefore be ignored; and (3) infection is lifelong due to auto-infection, which in terms of the other model^45^ is an “internal reproduction number” *R*_*I*_ > 1, which is consistent with the slow bounce-back of infection levels after treatment observed in the field.^17^ An important benefit of our simplifications is that they allow us to avoid several unidentifiable and highly uncertain parameters, while still allowing us to capture the effects of the associated uncertainty in the three transmission-related parameters of the model. We acknowledge that as a result of these simplifications, our model predicts that transmission is equally efficient at different endemicity levels, whereas in very low endemic settings, transmission could be relatively more efficient as it may take longer for hosts to build up an immune response due to lower exposure to incoming infections. However, this is mostly important for research questions related to interruption of transmission, which we do not consider here.

In our analyses, we captured uncertainty related to transmission conditions, the effectiveness of ivermectin, patterns in uptake of PC, and cost of procurement and delivery of ivermectin. However, we also adopted a number of simplifying assumptions that were not addressed in the main or sensitivity analysis and therefore require special attention. First, we simulated human demographic patterns that represent sub-Sahara Africa around the year 2000 (i.e., a high proportion of children in the population) and assumed that exposure and contribution to transmission is uniform over all ages, whereas the latter can also be considered to rise with age during the first ten years of life, as in some hookworm modelling studies.^20,21^ Both of these assumptions favour the cost-effectiveness of school-based PC compared to community-based PC, which means that in settings where adults make up a larger proportion of the population than assumed here or where exposure rises with age, school-based PC is even less cost-effective than predicted here and community-based PC should still be the preferred strategy.

Second, we assumed that the cure rate of ivermectin treatment is the same for all treated individuals, while in other soil-transmitted helminthiases, the cure rate depends on the hosts’ worm load ^18,19^. At most, this means that our estimates of cost-effectiveness are too optimistic for more highly endemic settings, where one might expect higher worm loads. However, that notion could be partially countered by the fact that *S. stercoralis* numbers are highly regulated by the host immune response and probably do not depend the host level of exposure but rather the rate at which a host auto-infects, which may not necessarily be correlated with endemicity. Also, community-based PC was predicted to be already highly cost-effective for settings in the second-lowest category for endemicity that we considered (baseline prevalence of 2% to 5% in school age children), meaning that, at most, the cost-effectiveness estimates for the higher endemicity categories are slightly worse than we predict, but still higher than for the 2-5% endemicity category (as cost-effectiveness improves with baseline endemicity). As such, we do not expect that this assumption of a fixed cure rate for ivermectin has biased our estimates in a way that would change our conclusions about the relative cost-effectiveness of different PC strategies.

Third, an important uncertainty in our calculations concerns the calculation of YLD due to infection, for which we conservatively assumed that only 25% of prevalent cases with infection experience symptoms. This assumption was based on the prevalence of symptoms among cases with diagnosed infection^26^ and the fact that the sensitivity of Baermann or stool culture is in the order of 50%.^11,27,28^ Therefore, we are ignoring potential YLDs in undiagnosed cases of infection, meaning that our estimates of cost-effectiveness are conservative.

However, as this equally affects our estimates for school-based and community-based PC, we do not expect that this uncertainty about the proportion of infected cases with symptoms affects our conclusions about community-based PC being the most cost-effective strategy.

Lastly, in the main analysis, we considered the simplest case where a PC program against strongyloidiasis would have to be set up and implemented as the sole NTD control program, whereas in reality, some sort of NTD control program may already be in place in many strongyloidiasis-endemic contexts. In the sensitivity analyses, we did however consider the optimal case where the delivery cost of school-based PC against strongyloidiasis might be considered zero because of a pre-existing PC program against STH. Still, in this case, community-based PC dominated school-based PC in terms of cost-effectiveness. Further, we did not consider potential averted costs related to hospitalisation of severe cases, although these were predicted to be relatively rare and probably would not have impacted cost estimates in a meaningful way. As such, for these simplifying assumptions, we do not expect them to have affected our conclusions about the superiority of community-based PC in terms of cost-effectiveness. Still, our estimates could be further refined with more detailed cost estimates, in particular by distinguishing between one-time start-up costs and annually returning costs of PC, and distinguishing types of cost that do and do not scale (or less so) with size of the target population.^46^

In conclusion, we predict that community-based PC is the most cost-effective PC strategy to control strongyloidiasis, being superior to school-based PC due to most of the infections and mortality occurring in adults. Its cost-effectiveness is well below or comparable to willingness to pay thresholds for low– and low-middle income countries. Even in settings where improved access to healthcare will reduce or already has reduced strongyloidiasis-related mortality, implementation community-based PC would still be cost-effective. A baseline prevalence threshold of 2% of infection in school age children, as measured by Baermann or stool culture, is in principle a suitable minimum for cost-effective implementation of community-based PC. Depending on the baseline endemicity and achieved PC coverage, implementing 3 up to 6 PC rounds will be cost-effective. For the purpose of identifying priority areas, it may be considered that the cost-effectiveness of PC strongly increases with baseline prevalence.

## Competing interests

NCL reports consulting fees from the World Health Organization related to guidelines on strongyloidiasis.

## Authors’ contributions

LEC conceived of the study, designed and performed the analyses, programmed the mathematical model, visualised and interpreted the results, and drafted the manuscript. NCL investigated and formulated appropriate disability weights for strongyloidiasis sequelae, interpreted the results, and revised the manuscript. SJdV contributed to the design of the analyses, interpreted the results, and revised the manuscript.

## Funding

The research described here was performed as part of an Agreement for Performance of Work between Erasmus MC, University Medical Center Rotterdam, The Netherlands and the World Health Organization, Geneva, Switzerland under WHO Registration 2023/1339288-1 and WHO Purchase Order 203102918-1 (awarded to LEC).

## Data Availability

All data used in this study originate from the public domain (i.e., estimates from published literature) or are available as open-source software. The output data from the simulation model and the code that generated it are available on request and will be made readily and publicly available for the peer-reviewed version of the paper.

https://gitlab.com/luccoffeng/strongysim

## Acknowledgements

The authors gratefully acknowledge scientific experts Antonio Montresor, Dora Buonfrate, Francesca Tamarozzi, Jennifer Keiser, and Alejandro Krolewiecki for their advice on unknown and uncertain aspects of strongyloidiasis biology, diagnostics, and treatment. The authors are further grateful to Lorenzo Moja (methodologist, WHO Geneva) for his review of the methods used in this study.

## Supplemental information

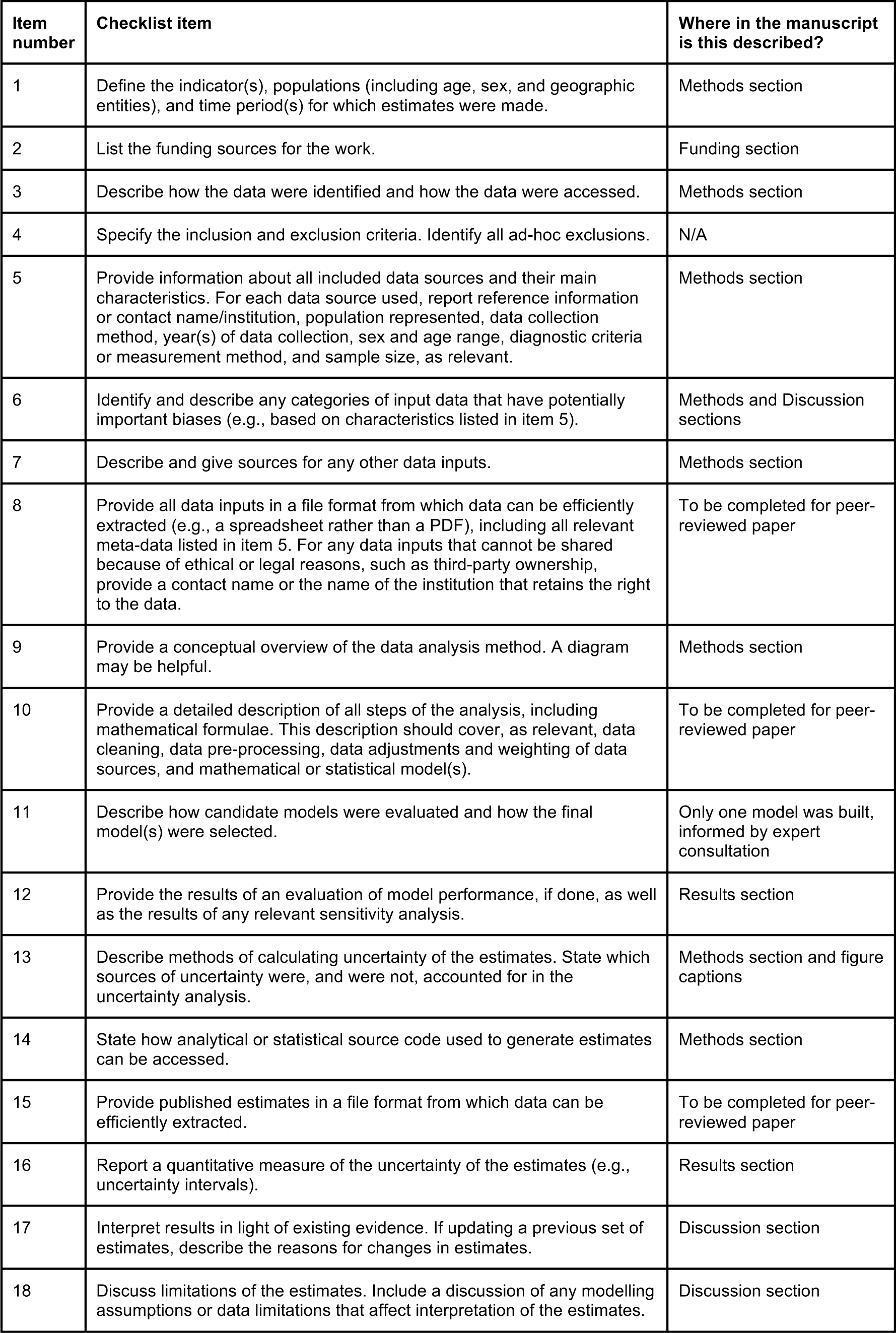
Guidelines for Accurate and Transparent Health Estimates Reporting (GATHER)^41^.

| Item number | Checklist item | Where in the manuscript is this described? |
| --- | --- | --- |
| 1 | Define the indicator(s), populations (including age, sex, and geographic entities), and time period(s) for which estimates were made. | Methods section |
| 2 | List the funding sources for the work. | Funding section |
| 3 | Describe how the data were identified and how the data were accessed. | Methods section |
| 4 | Specify the inclusion and exclusion criteria. Identify all ad-hoc exclusions. | N/A |
| 5 | Provide information about all included data sources and their main characteristics. For each data source used, report reference information or contact name/institution, population represented, data collection method, year(s) of data collection, sex and age range, diagnostic criteria or measurement method, and sample size, as relevant. | Methods section |
| 6 | Identify and describe any categories of input data that have potentially important biases (e.g., based on characteristics listed in item 5). | Methods and Discussion sections |
| 7 | Describe and give sources for any other data inputs. | Methods section |
| 8 | Provide all data inputs in a file format from which data can be efficiently extracted (e.g., a spreadsheet rather than a PDF), including all relevant meta-data listed in item 5. For any data inputs that cannot be shared because of ethical or legal reasons, such as third-party ownership, provide a contact name or the name of the institution that retains the right to the data. | To be completed for peer-reviewed paper |
| 9 | Provide a conceptual overview of the data analysis method. A diagram may be helpful. | Methods section |
| 10 | Provide a detailed description of all steps of the analysis, including mathematical formulae. This description should cover, as relevant, data cleaning, data pre-processing, data adjustments and weighting of data sources, and mathematical or statistical model(s). | To be completed for peer-reviewed paper |
| 11 | Describe how candidate models were evaluated and how the final model(s) were selected. | Only one model was built, informed by expert consultation |
| 12 | Provide the results of an evaluation of model performance, if done, as well as the results of any relevant sensitivity analysis. | Results section |
| 13 | Describe methods of calculating uncertainty of the estimates. State which sources of uncertainty were, and were not, accounted for in the uncertainty analysis. | Methods section and figure captions |
| 14 | State how analytical or statistical source code used to generate estimates can be accessed. | Methods section |
| 15 | Provide published estimates in a file format from which data can be efficiently extracted. | To be completed for peer-reviewed paper |
| 16 | Report a quantitative measure of the uncertainty of the estimates (e.g., uncertainty intervals). | Results section |
| 17 | Interpret results in light of existing evidence. If updating a previous set of estimates, describe the reasons for changes in estimates. | Discussion section |
| 18 | Discuss limitations of the estimates. Include a discussion of any modelling assumptions or data limitations that affect interpretation of the estimates. | Discussion section |

Policy-Relevant Items for Reporting Models in Epidemiology of Neglected Tropical Diseases (PRIME-NTD)^42^.

| Principle | What has been done to satisfy the principle? | Where in the manuscript is this described? |
| --- | --- | --- |
| Stakeholder engagement | This work was commissioned by the stakeholder, the World Health Organization (WHO), which provided the authors with access to the WHO dossier on strongyloidiasis and meetings with the experts on the WHO Guideline Development Group for strongyloidiasis control. | Methods section, subsection "Role of the funding source" |
| Complete model documentation | The model was developed as a fully documented open-source R package, including help files and an example of how to run the model. | Methods section |
| Complete description of data used | The literature data sources used to inform parameter values of the model used in this work are described and cited in the methods section. | Methods section |
| Communicating uncertainty | Where relevant and useful, we present uncertainty as 95%-confidence interval, explaining in captions what types of uncertainty are captured. | Methods and results sections |
| Testable model outcomes | Expected trends in infection, and importantly, the speed of bounce-back of population infection levels between treatment rounds are visualised. These trends will be important to confirm with field experiments. | Figure 3 |

## Supplemental Figures

**Supplemental Figure A.**
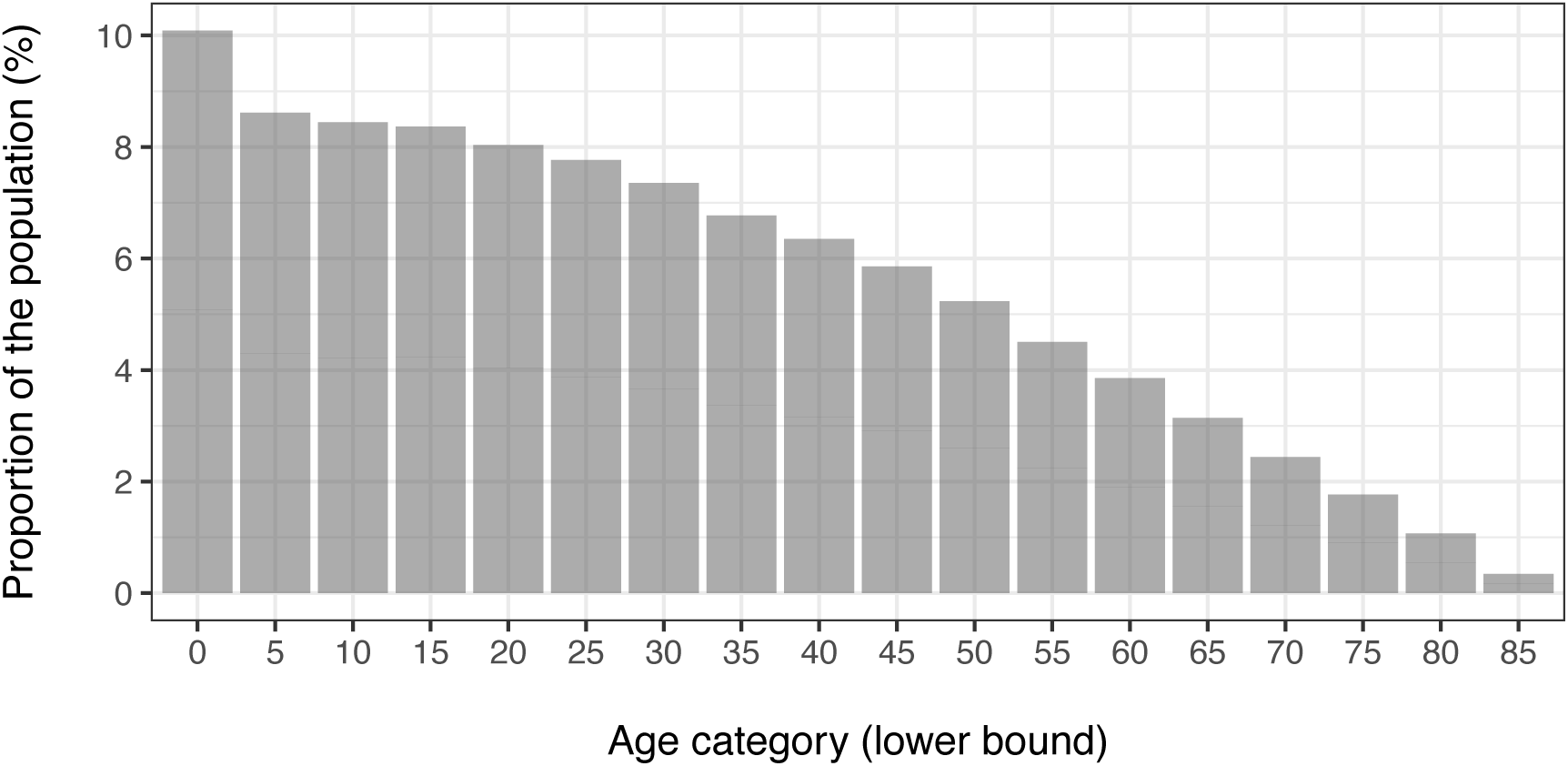
Age distribution of simulated populations, using demographic and mortality data from sub-Sahara Africa in 2000. In countries that have already (partly) gone through the first and/or second demographic transition (first = reduced child mortality; second = reduced birth rate), populations may consist of relatively more adults than we simulate here. As such, the demography we simulate somewhat favours school-based deworming as a relatively larger proportion of the population is captured by such a strategy.

**Supplemental Figure B.**
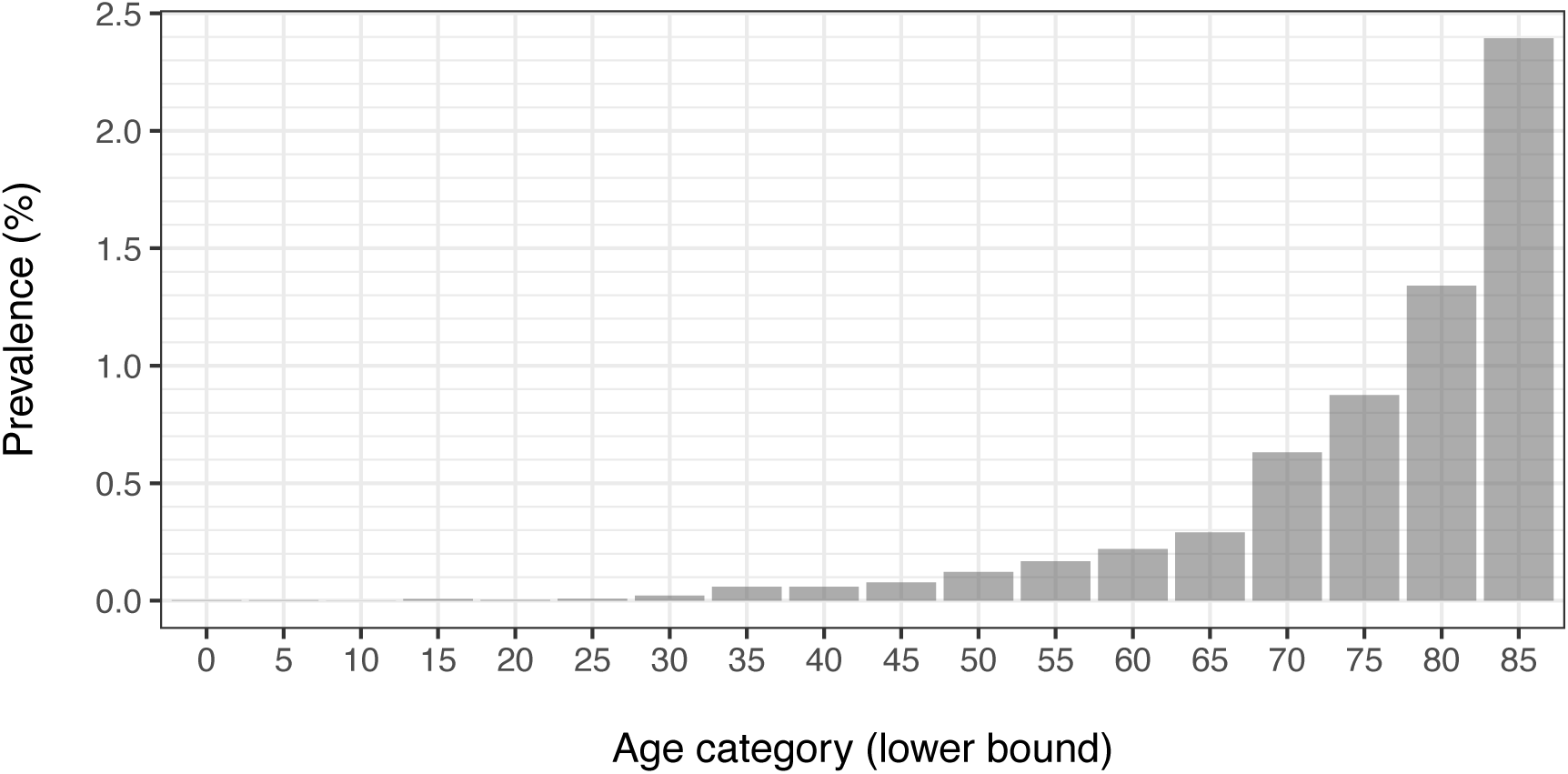
Simulated prevalence of immune-compromised states in a strongyloidiasis-free population. The prevalence pattern is result of the assumption that the average incidence of immune-compromised states per life-year is such that the 80-year-life-time risk is 2.5% and that the immune-compromised state develops at 95% of the total strongyloidiasis-free life expectancy. Prevalence for ages 80+ was not exactly 2.5% due to stochastic variation, which was considerable for higher age groups due to a relatively low number of individuals. The overall prevalence of immune-compromised states in the plotted population was 0.1%. The figure is based on a one-time simulation of a population of 500 thousand people.

**Supplemental Figure C.**
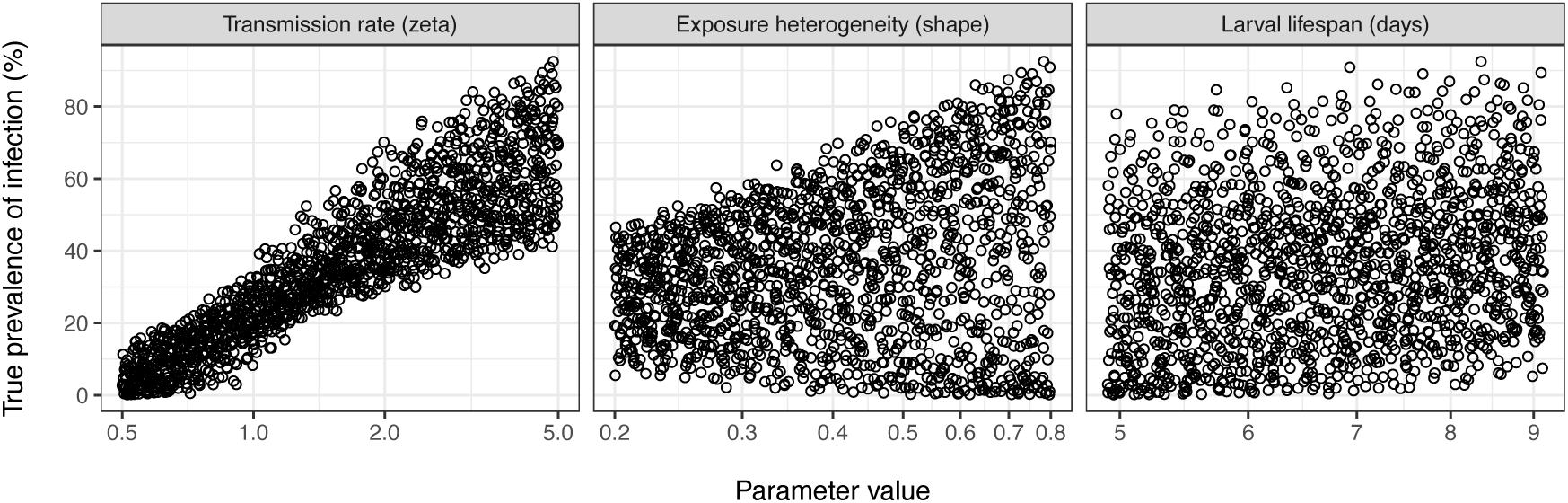
Simulated baseline prevalence of infection as a function of transmission conditions. Each point represents one of 1,500 random transmission conditions. Transmission conditions were defined in terms of the overall transmission rate (left), the level of exposure heterogeneity (middle; lower parameter values represent higher heterogeneity), and the average lifespan of free-living infective larvae in the environment (right). Plotted prevalences represent prevalence in the entire population as if measured by a 100% sensitive and 100% specific diagnostic tool.

**Supplemental Figure D.**
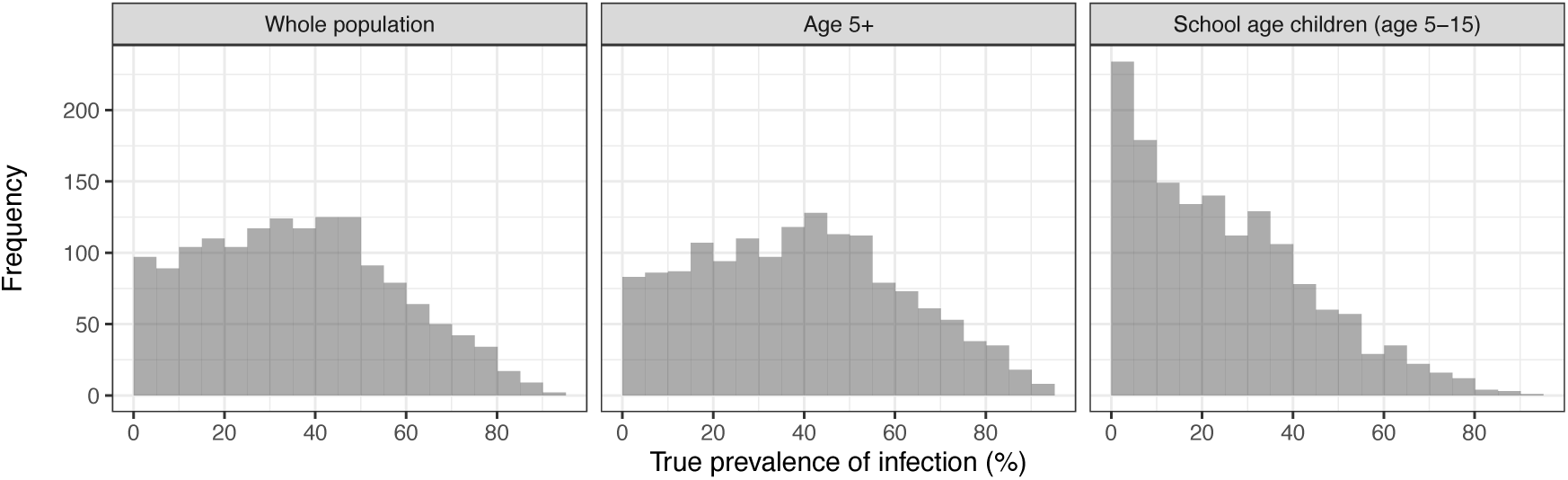
Histogram of simulated baseline prevalence of infection by age group. Each histogram represents 1,500 random transmission conditions (i.e., all bars add up to 1,500). Simulated baseline prevalences represent prevalence as if measured by a 100% sensitive and 100% specific diagnostic tool, and are shown for three age groups.

**Supplemental Figure E.**
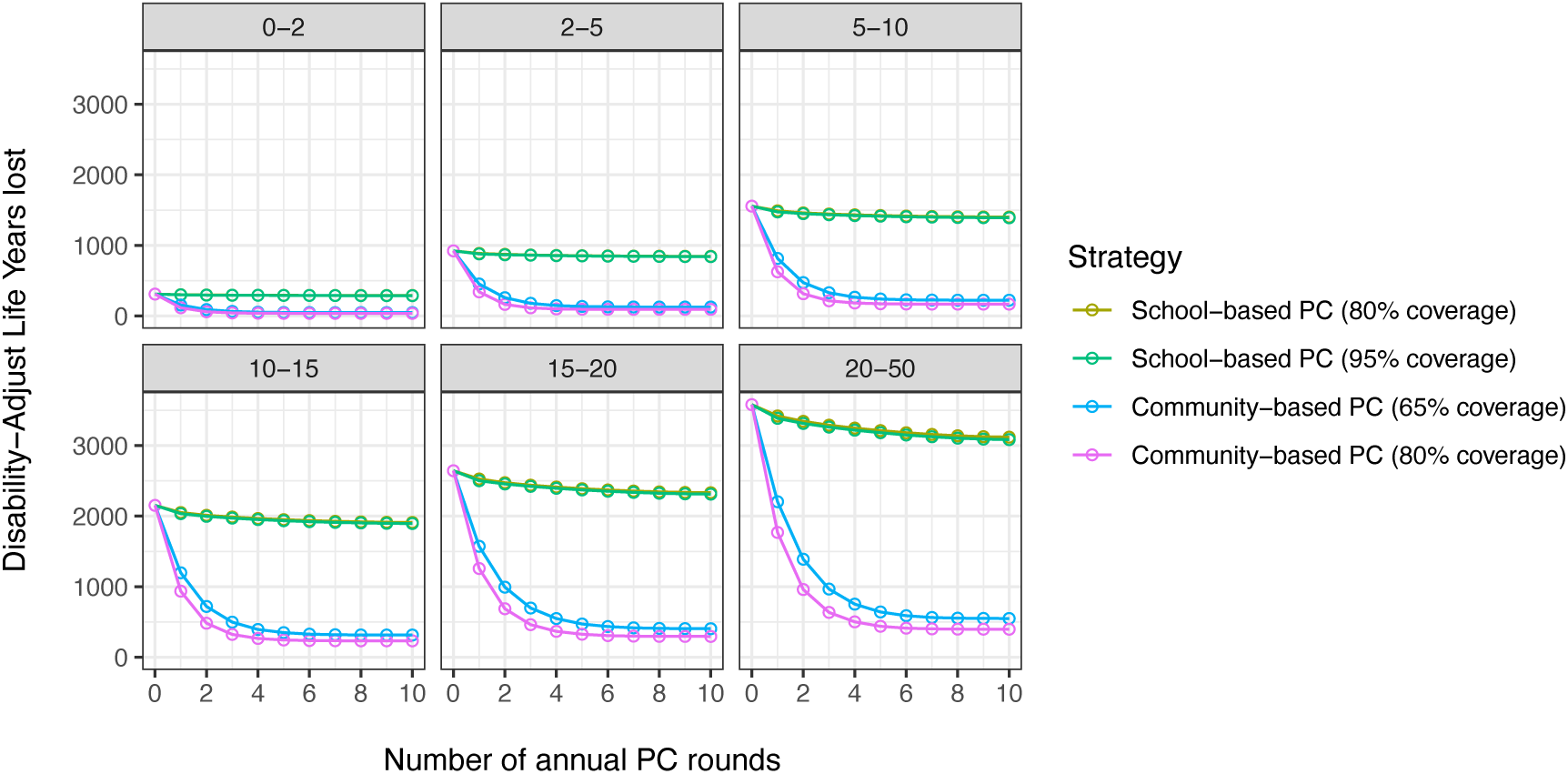
Disability-adjusted life-years (DALYs) lost due to strongyloidiasis over 10 years under different scenarios for the number of annual preventive chemotherapy (PC) rounds. Panels represents different endemicity scenarios; panel labels indicate the baseline prevalence of infection (%) in school age children, as measured by a coprological test with 50% sensitivity and 100% specificity. Colours indicate different PC strategies in terms of target population and achieved coverage.

**Supplemental Figure F.**
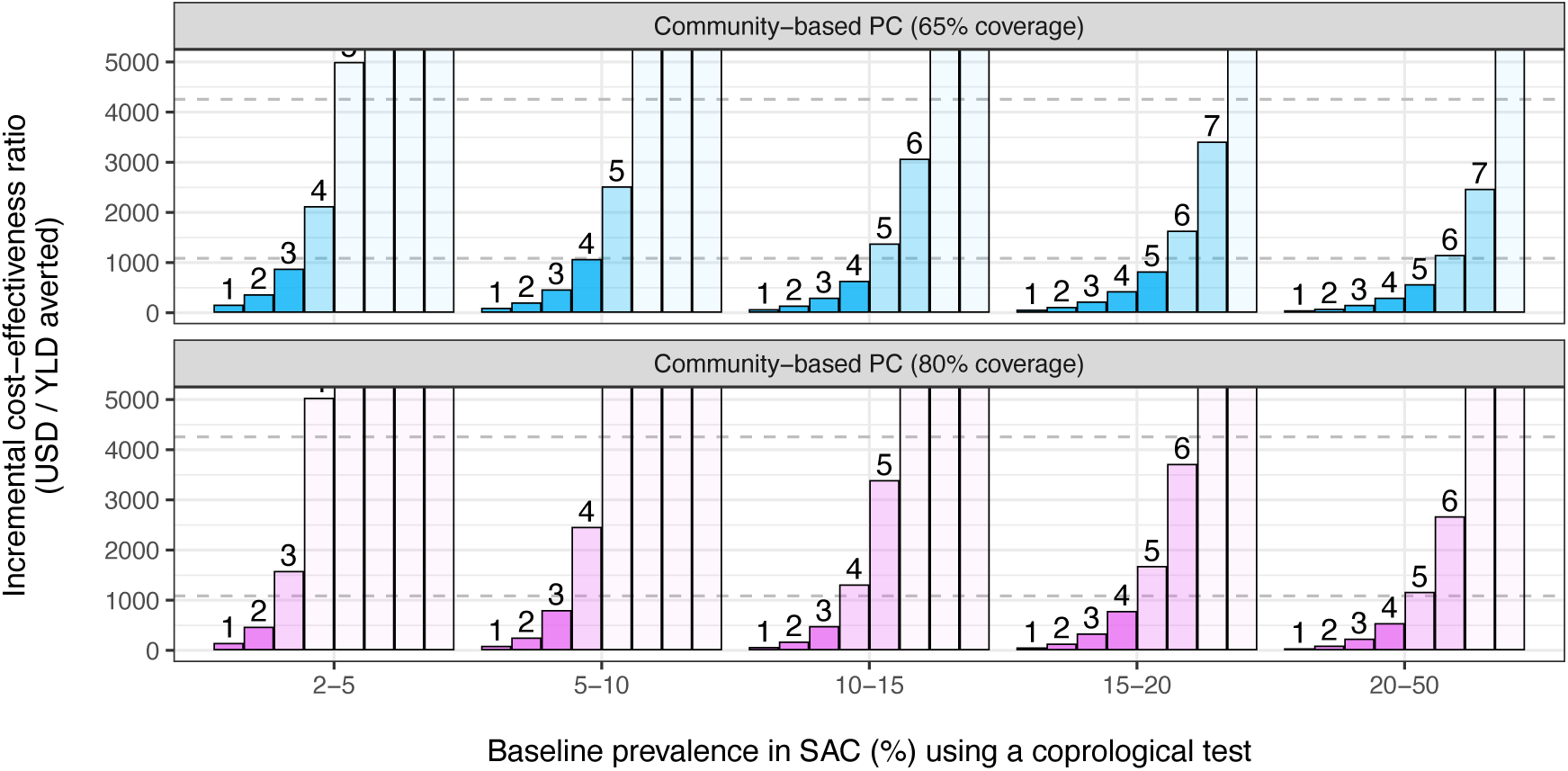
Incremental cost-effectiveness ratios (ICERs), based on years lived with disability or YLD), for increasing number of annual rounds of community-based preventive chemotherapy (PC). ICERs were calculated based on the cost of PC and the total number of disability-adjusted life-years lost over a 10-year period, applying a 3% discounting rate to both cost and effects. The number over each bar indicates the number of PC rounds, where the colour of bars indicates whether the ICER for that number of PC rounds was under the average gross domestic product (GDP) for low-income countries (US$1,085, lower dashed line) or under the average GDP of lower middle-income countries (US$4,255, upper dashed line), or over.^39^ ICERs greater than US$5,000 are capped at the top of each panel. Note that ICERs may be underestimated for the shortest PC program durations. This is because an average cost per treatment (calculated for a 10-year period) was applied to all PC program durations, not considering a potential concentration of costs at the start of PC programs. This also means that the ICERs for longer program durations may be lower than presented here.

## Supplemental Tables

**Supplemental Table A.** Model-predicted number of life-years lived with infection and the absolute number of hyperinfections per 100,000 population over 10 years, and the associate health burden. Twenty percent of cases of hyperinfection (95%-CI: 10–30%) were assumed to be successfully diagnosed and treated; all other cases of hyperinfection were assumed to die, losing 5% of their life-expectancy. All estimates of life-years, cases, and burden include an annual discounting rate of 3%. Point estimates represent medians over repeated simulated populations. Numbers in brackets represent the central 95%-confidence intervals that capture uncertainty about costs, disability weights, the fraction of the hyperinfections that are prevented from dying via regular routine healthcare, and stochastic variation in a population of 100,000 people.

| Baseline prevalence in SAC (%)* | Strategy | Coverage of PC | Life-years with infection (x1,000) | Number of hyperinfections | YLLs lost | YLDs lost | DALYs lost |
| --- | --- | --- | --- | --- | --- | --- | --- |
| 0-2 | No PC | - | 45 (28-62) | 32 (17-49) | 84 (46-135) | 219 (109-392) | 304 (168-501) |
|  | School-based PC | 80% | 41 (26-56) | 32 (18-49) | 84 (47-134) | 200 (100-354) | 285 (158-465) |
|  |  | 95% | 40 (26-55) | 31 (17-49) | 83 (47-132) | 198 (99-353) | 283 (157-459) |
|  | Community-based PC | 65% | 4 (3-6) | 9 (4-16) | 24 (10-46) | 22 (11-38) | 46 (24-75) |
|  |  | 80% | 3 (2-4) | 8 (3-15) | 21 (7-41) | 14 (7-25) | 36 (18-60) |
| 2-5 | No PC | - | 137 (120-155) | 87 (67-109) | 233 (172-305) | 671 (401-1,080) | 903 (635-1,326) |
|  | School-based PC | 80% | 122 (107-137) | 86 (67-107) | 233 (174-305) | 596 (355-953) | 829 (581-1,206) |
|  |  | 95% | 121 (106-136) | 87 (66-108) | 233 (171-307) | 592 (352-949) | 826 (583-1,201) |
|  | Community-based PC | 65% | 14 (12-16) | 21 (12-30) | 56 (32-83) | 69 (42-109) | 125 (86-173) |
|  |  | 80% | 9 (8-10) | 18 (10-26) | 48 (26-74) | 43 (26-69) | 92 (61-128) |
| 5-10 | No PC | - | 238 (216-260) | 137 (114-164) | 366 (292-456) | 1,175 (682-1,815) | 1,537 (1,048-2,206) |
|  | School-based PC | 80% | 207 (189-226) | 135 (112-161) | 362 (290-452) | 1,024 (592-1,589) | 1,383 (957-1,970) |
|  |  | 95% | 206 (187-224) | 135 (112-160) | 362 (289-450) | 1,015 (587-1,572) | 1,374 (955-1,952) |
|  | Community-based PC | 65% | 25 (23-28) | 36 (26-50) | 96 (65-134) | 125 (72-192) | 221 (159-305) |
|  |  | 80% | 16 (14-17) | 33 (22-45) | 87 (57-121) | 79 (46-122) | 166 (121-223) |
| 10-15 | No PC | - | 337 (316-358) | 178 (149-207) | 468 (379-573) | 1,663 (969-2,613) | 2,117 (1,437-3,066) |
|  | School-based PC | 80% | 289 (271-307) | 176 (147-205) | 465 (376-570) | 1,426 (832-2,243) | 1,884 (1,295-2,683) |
|  |  | 95% | 286 (268-304) | 176 (147-206) | 466 (374-571) | 1,412 (825-2,220) | 1,869 (1,283-2,660) |
|  | Community-based PC | 65% | 37 (35-39) | 49 (36-65) | 128 (90-176) | 183 (108-286) | 313 (229-419) |
|  |  | 80% | 24 (22-25) | 44 (30-58) | 113 (76-158) | 117 (69-182) | 232 (169-305) |
| 15-20 | No PC | - | 420 (397-441) | 208 (181-239) | 546 (442-663) | 2,068 (1,203-3,208) | 2,614 (1,764-3,767) |
|  | School-based PC | 80% | 358 (339-375) | 206 (178-237) | 544 (441-662) | 1,762 (1,024-2,729) | 2,304 (1,591-3,292) |
|  |  | 95% | 353 (335-371) | 206 (178-236) | 544 (444-661) | 1,743 (1,012-2,697) | 2,285 (1,580-3,264) |
|  | Community-based PC | 65% | 50 (47-52) | 61 (45-75) | 156 (111-202) | 244 (144-383) | 402 (297-549) |
|  |  | 80% | 31 (30-33) | 53 (38-67) | 136 (97-180) | 155 (92-242) | 293 (218-389) |
| 20-50 | No PC | - | 577 (532-627) | 267 (233-303) | 695 (571-838) | 2,859 (1,658-4,330) | 3,553 (2,343-5,036) |
|  | School-based PC | 80% | 485 (449-525) | 265 (231-301) | 693 (570-835) | 2,407 (1,398-3,646) | 3,091 (2,082-4,366) |
|  |  | 95% | 478 (443-516) | 264 (230-300) | 693 (571-834) | 2,371 (1,377-3,588) | 3,059 (2,058-4,316) |
|  | Community-based PC | 65% | 70 (63-78) | 78 (60-96) | 195 (147-262) | 347 (202-534) | 543 (387-742) |
|  |  | 80% | 45 (41-50) | 67 (50-84) | 169 (126-225) | 223 (130-343) | 394 (287-530) |

\* Baseline coprovalence in school age children (SAC), assuming 50% sensitivity of the diagnostic test used.
PC = preventive chemotherapy; YLL = Years of Life Lost; YLD = Years Lived with Disability; DALY = Disability-Adjusted Life-Years.

**Supplemental Table B.** Cost-effectiveness and incremental cost-effectiveness ratios of different 10-year preventive chemotherapy (PC) strategies, based on averted Years Lived with Disability (YLDs). Note that costs-effectiveness and ICER estimates are expressed in US dollars (not thousands). Cost of distribution of ivermectin was assumed to be as estimated for preventive chemotherapy against soil-transmitted helminths in Dak Lak province, Vietnam.^33^ The cost-effectiveness and incremental cost-effectiveness ratios in terms of solely YLDs reflect the assumption that there is zero strongyloidiasis-related mortality. Point estimates represent medians over repeated simulated populations of 100,000 people. Numbers in brackets represent the central 95%-confidence intervals that capture uncertainty about costs, disability weights, and stochastic variation in a population of 100,000 people.

| Baseline prevalence in SAC (%)* | PC Strategy | PC Coverage | Cost-effectiveness (USD per YLD averted) |  | Incremental cost-effectiveness ratio (USD per additional YLD averted) ** |  |
| --- | --- | --- | --- | --- | --- | --- |
| 0-2 | School-based PC | 80% | 4,504 | (2,352-9,694) | - |  |
|  |  | 95% | 4,755 | (2,481-10,140) | - |  |
|  | Community-based PC | 65% | 2,440 | (1,365-4,932) | 2,440 | (1,365-4,932) |
|  |  | 80% | 2,698 | (1,503-5,446) | 9,039 | (4,852-19,230) |
| 2-5 | School-based PC | 80% | 1,199 | (709-2,157) | - |  |
|  |  | 95% | 1,253 | (734-2,279) | - |  |
|  | Community-based PC | 65% | 797 | (495-1,362) | 797 | (495-1,362) |
|  |  | 80% | 879 | (534-1,499) | 2,805 | (1,627-4,914) |
| 5-10 | School-based PC | 80% | 596 | (362-1,073) | - |  |
|  |  | 95% | 621 | (380-1,126) | - |  |
|  | Community-based PC | 65% | 460 | (293-782) | 460 | (293-782) |
|  |  | 80% | 507 | (319-864) | 1,575 | (893-2,729) |
| 10-15 | School-based PC | 80% | 386 | (237-680) | - |  |
|  |  | 95% | 401 | (245-695) | - |  |
|  | Community-based PC | 65% | 328 | (207-555) | 328 | (207-555) |
|  |  | 80% | 360 | (226-605) | 1,082 | (616-1,841) |
| 15-20 | School-based PC | 80% | 298 | (184-522) | - |  |
|  |  | 95% | 308 | (189-535) | - |  |
|  | Community-based PC | 65% | 265 | (166-448) | 265 | (166-448) |
|  |  | 80% | 291 | (183-488) | 804 | (464-1,358) |
| 20-50 | School-based PC | 80% | 200 | (121-357) | - |  |
|  |  | 95% | 205 | (125-368) | - |  |
|  | Community-based PC | 65% | 193 | (121-339) | 193 | (121-339) |
|  |  | 80% | 212 | (133-372) | 581 | (330-1,001) |

\* Baseline coprovalence in school age children (SAC), assuming 50% sensitivity and 100% specificity of the diagnostic test used.
\*\* Dashes indicate strategies that are extended dominated, meaning that although they did provide additional benefits (DALYs or YLDs averted) but at a higher cost per additional unit of effect than other strategies.

